# Modelling outbreak response strategies for preventing spread of emergent *Neisseria gonorrhoeae* strains in men who have sex with men in Australia

**DOI:** 10.1101/2021.04.30.21256375

**Authors:** Qibin Duan, Chris Carmody, Basil Donovan, Rebecca J Guy, Ben B Hui, John M Kaldor, Monica M Lahra, Matthew G Law, David A Lewis, Michael Maley, Skye McGregor, Anna McNulty, Christine Selvey, David J Templeton, David M Whiley, David G Regan, James G Wood

**Author notes:** Corresponding authors: James G Wood, School of Population Health, UNSW Sydney, Sydney, NSW 2052, Australia., David G Regan, The Kirby Institute, UNSW Sydney, Sydney, NSW 2052, Australia.

## Abstract

The ability to treat gonorrhoea with current first-line drugs is threatened by the global spread of extensively drug resistant (XDR) *Neisseria gonorrhoeae* (NG) strains. In Australia, urban transmission is high among men who have sex with men (MSM) and emergence of an imported XDR NG strain in this population could result in an epidemic that would be difficult and costly to control. An individual-based, anatomical site-specific mathematical model of NG transmission among Australian MSM was developed and used to evaluate the potential for elimination of an emergent XDR NG strain under a range of case-based and population-based test-and-treat strategies. When applied upon detection of the imported strain, these strategies enhanced the probability of elimination and reduced the outbreak size compared with current practice. The most effective strategies combined testing targeted at regular and casual partners with increased rates of population testing. However, even with the most effective strategies, outbreaks could persist for up to 2 years post-detection. Our simulations suggest that local elimination of emergent NG XDR strains can be achieved with high probability using combined case-based and population-based test-and-treat strategies. These strategies may be an effective means of preserving current treatments in the event of wider XDR NG emergence.

**Author Summary:** In most high-income settings, gonorrhoea transmission is endemic among men who have sex with men (MSM). While gonorrhoea remains readily treatable, there are major concerns about further resistance due to recent reports of treatment failure with first-line therapy and limited remaining treatment options. Here we investigated the potential for trace and treat response strategies to eliminate such strains before their prevalence reaches a level requiring a shift to new first line therapies. Rather than directly consider resistance, we explore the mitigating effect of various test and trace measures on outbreaks of a generic imported strain which remains treatable. This is done within a realistic mathematical model of spread in an MSM community that captures cases, anatomical sites of infection and contacts at an individual level, calibrated to Australian epidemiology. The results indicate that strategies such as partner tracing and treatment in combination with elevated asymptomatic community testing are highly effective in mitigating outbreaks but can take up to 2 years to achieve elimination. As there are currently no clear alternatives of proven efficacy and safety to replace ceftriaxone in first-line therapy, these promising results suggest potential for use of these outbreak response strategies to enable continuation of current treatment recommendations.

## Introduction

Gonorrhoea is a sexually transmissible infection caused by the bacterium *Neisseria gonorrhoeae* (NG). Antibiotics have, for many decades, provided effective treatment for gonorrhoea. However, resistance to several classes of antimicrobial agents has emerged, rendering many previously effective drugs ineffective (1). Ceftriaxone is now recommended in most countries as the backbone of first line therapy for gonorrhoea. Since 2009, reports of NG strains exhibiting resistance to ceftriaxone have generated substantial concern in national and global public health agencies (2, 3). These reports were initially sporadic but by 2017 evidence emerged of sustained spread of ceftriaxone-resistant strains harbouring a novel resistance mechanism in the form of a *penA* type 60.001 allele (4). Since then, a further extensively drug resistant (XDR) strain harbouring the *penA* 60.001 allele as well as high-level resistance to azithromycin has been reported in Australia, the UK and continental Europe (5-8). With the possible exception of spectinomycin and gentamicin (9), there are no alternative drugs of proven safety and efficacy currently available for routine treatment of anogenital gonorrhoea. Novel antimicrobials are being trialled but are yet to be comprehensively assessed (10, 11).

Without effective treatment and/or surveillance, there is the potential for rapid spread of resistance within populations as highlighted in South Australia in 2016, where azithromycin resistance rose from 5% to 30% of isolates within just 12 weeks, before dropping back to 12% in 2017 (12). Rapid rises in ciprofloxacin resistance in New South Wales, Australia between 1991 and 1997 (13) and in South Africa in 2003 (14) have also been reported. These examples, combined with the recently reported cases of XDR NG, suggest that larger outbreaks of these or similar XDR strains are imminent, potentially arriving in Australia via repeated importation from the Asia-Pacific region as has been observed previously (13, 15).

The prevalence of gonorrhoea in Australia is highest in remote Aboriginal and Torres Strait Islander populations and among men who have sex with men (MSM) in metropolitan centres. Although to date most reported cases of XDR NG have involved heterosexual contact (5, 7, 8, 16), we have chosen to focus on MSM due to the high incidence of gonorrhoea in this population, frequent sexual contact when travelling overseas, and evidence of the importance of oropharyngeal NG in driving transmission in this population (17, 18). These factors together suggest that establishment of an XDR NG strain within an MSM population could spark a rapidly expanding gonorrhoea epidemic that would be difficult to contain (19).

Without an effective gonococcal vaccine, developing appropriate public health responses to identify cases, reduce onward transmission, and maintain effective treatment for NG infections, are now key strategic priorities. However, beyond further changes to recommended antibiotic regimens, there is scant published evidence to inform the design of such responses. In this study we use a mathematical model of gonorrhoea transmission in an urban Australian MSM population to evaluate the potential impact of current and alternative surveillance and control strategies on the persistence of imported/emergent NG strains. Note that this study mainly focuses on outbreaks attributable to such imported strain.

## Results

### Model Simulation and Calibration

An individual-based model was developed that captures the dynamic formation and dissolution of sexual partnerships in a population of 10,000 MSM, sexual acts within partnerships, and the transmission of NG between 3 anatomical sites: urethra, oropharynx, anorectum. The natural history of gonorrhoea is captured in a Susceptible-Exposed-Infectious-Recovered (SEIRS) framework. Parameter values relating to gonorrhoea natural history have been derived, where possible, from published literature and are listed in Table 1. In each daily simulation cycle (illustrated in Figure 1) transmission events are tracked and the infectious status of all individuals updated. Events relating to natural progression of infection, testing, treatment of infection and entry/exit of individuals from the sexually active population are then processed before concluding each simulation cycle with partnership formation and dissolution.

**Table 1.** Gonorrhoea infection parameters and prevalence targets for model calibration.

| Parameter description | Value | Source |
| --- | --- | --- |
| *Proportion of infections that are symptomatic by anatomical site* |  |  |
| Oropharyngeal infection | 0 | Oropharyngeal gonorrhoea is rarely associated with symptoms (42) |
| Urethral infection | 90% | (43, 44) |
| Anorectal infection | 12% | (45) |
| *Average duration of infection stages (range)* |  |  |
| Oropharyngeal infection | 84 days (70,138) | Sampled from $\Gamma(201,0.4)$ within the specified range (46, 47) |
| Urethral infection | 84 days (70, 140) | Sampled from $\Gamma(206,0.4)$ within the specified range (17, 18) |
| Anorectal infection | 343 days (336,361) | Sampled from $\Gamma(3702,0.1)$ within the specified range (47) |
| Time from onset of anorectal/urethral symptoms to treatment | 3 days (1,7) | Sampled from $\Gamma(3,0.86)$ within the specified range (48) |
| Incubation | 4 days (2,10) | Sampled from $\text{Exp}(4)$ within the specified range (49) |
| Exposed | 3.6 days (1,9) | Sampled from $U(1, \text{length of incubation period})$ within the specified range |
| Immunity | 3.5 days (1,7) | Assumption based on (50, 51) |
| *Prevalence targets for model calibration (95% CI)* |  |  |
| Oropharynx | 8.6% (7.7-9.5) | Based on (18) and comparable |
| Anorectum | 8.3% 7.4-9.1) | with (52) |
| Urethra | 0.26% (0.04-0.35) | Based on (18) |

**Figure 1.**
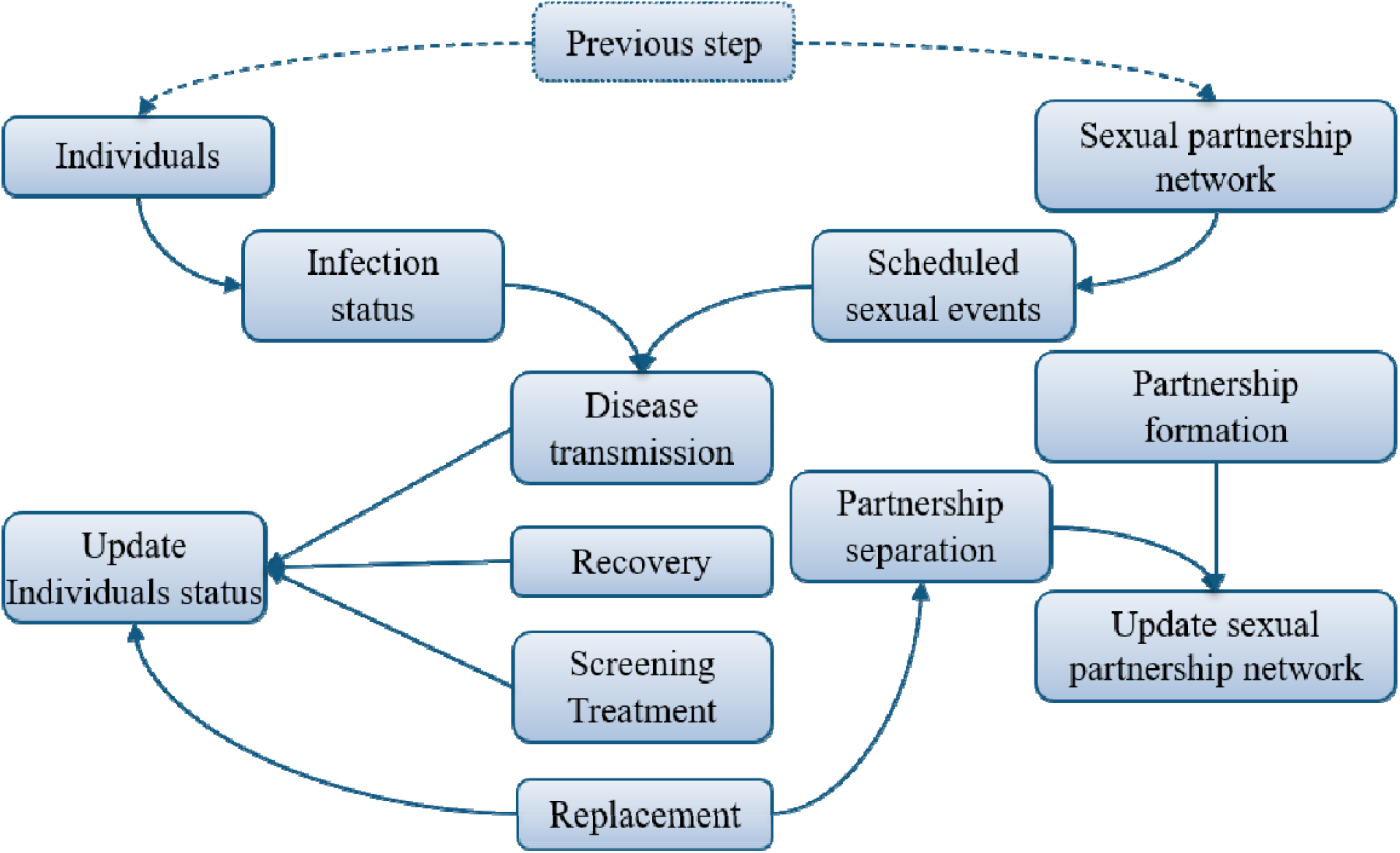
Schematic illustration of sequence of events that occur in a single simulation cycle of the individual-based model.

The model was calibrated to estimated anatomical site-specific NG prevalence in a hypothetical community sample as reported in Zhang *et al*. (18): oropharynx 8.6%; anorectum 8.3%; urethra 0.26% (Table 1). Comparison of 50 simulations from the calibrated model to prevalence targets is shown in Figure 2. Equilibrium is reached at approximately three years, with the site-specific prevalence curves then fluctuating around the target values. Figures A1 and A2 (Technical Appendix) provide validation that the characteristics of the model-generated sexual contact network are consistent with data reported in the GCPS Sydney 2018 (20) and the HIM Study (21).

**Figure 2.**
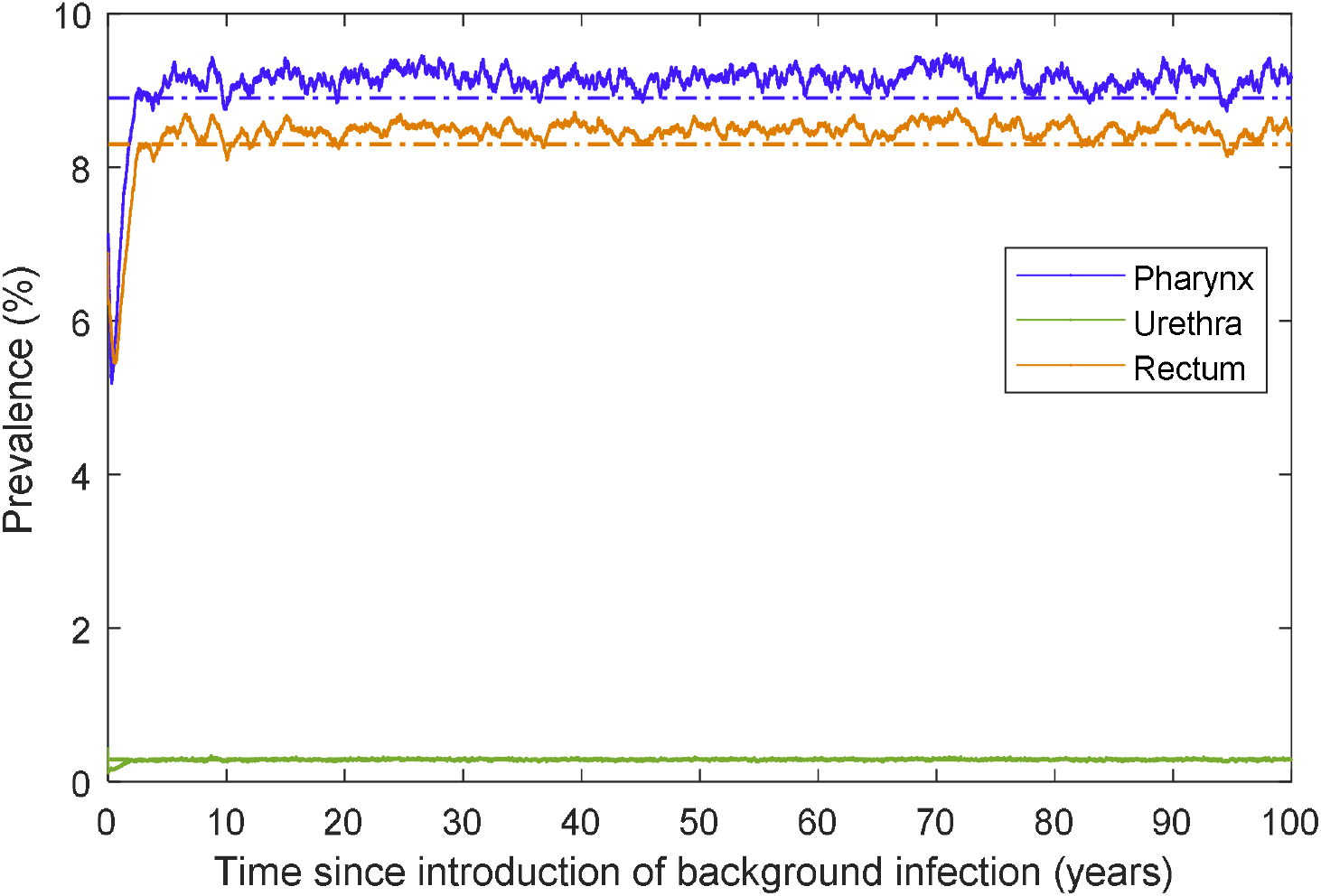
Average daily model-generated site-specific prevalence over 50 simulations (solid lines) and calibration targets (dashed lines). Each simulation was run for 100 years using the per-act transmission probabilities obtained through the calibration process and daily site-specific prevalence was averaged over the 50 simulations.

### Impact of Outbreak Response Strategies

#### Outbreak Response Strategies

The calibrated model is then used to assess the effectiveness of intervention strategies in controlling strain importations. To simulate importation/emergence, we first select an infection site based on relative site-specific incidence rates reported in Callander et. al. (22), yielding an oro-pharynx: urethra: rectum imported case ratio of 47:23:30. An imported gonococcal strain is then seeded in a randomly selected individual, already infected with the endemic strain, at the selected anatomical site. The endemic strain is removed immediately prior to strain importation, with simulations focusing on outbreaks related to the imported strain. We assume the imported strain is detectable and treatable. Note that the imported strain is not particularly resistant and can be treated with current treatment or alternatives.

We consider two levels of testing coverage (Table A6, Technical Appendix): current testing (CT) reflects testing coverage as reported in GCPS Sydney 2018 (20) and ACCESS (23); recommended testing (RT) is based on the 2016 Australian STI Management Guidelines (24) according to which all MSM should test at least once per year and those with 20+ sexual partners per year should test every three months. We also provide results with testing rates according to the 2019 Sexually Transmissible Infections in Gay Men Action Group (STIGMA) guidelines 2019 (25), which recommends 3 monthly testing for most sexually active MSM not in monogamous relationships, as a sensitivity analysis in the Technical Appendix. We assume testing occurs at all anatomical sites simultaneously with 100% sensitivity, 100% treatment efficacy in individuals who test positive for gonorrhoea infection, and clearance of viable gonococci from all anatomical sites within 1 day. An alternative scenario assuming 95% test sensitivity, 95% treatment efficacy, and a 7-day clearance delay for asymptomatic infection is presented in the Technical Appendix.

Further, we examine the effect of combining different levels of sexual contact-based testing and treatment in combination with current/recommended screening. The Australian Contact Tracing Guidelines (26) advise that recent (last 2 months) sexual contacts of index patients with gonorrhoea should be offered testing and treatment to minimise reinfection and onward transmission. We consider provision of testing/treatment to 80% or 100% of current regular partners (PTTR80 and PTTR) and four additional strategies, whereby 20%, 30%, 40% or 50% of casual partners in the last 2 months are tested/treated (PTT_RC20_, PTT_RC30_, etc.) as summarised in Table 2.

**Table 2.**
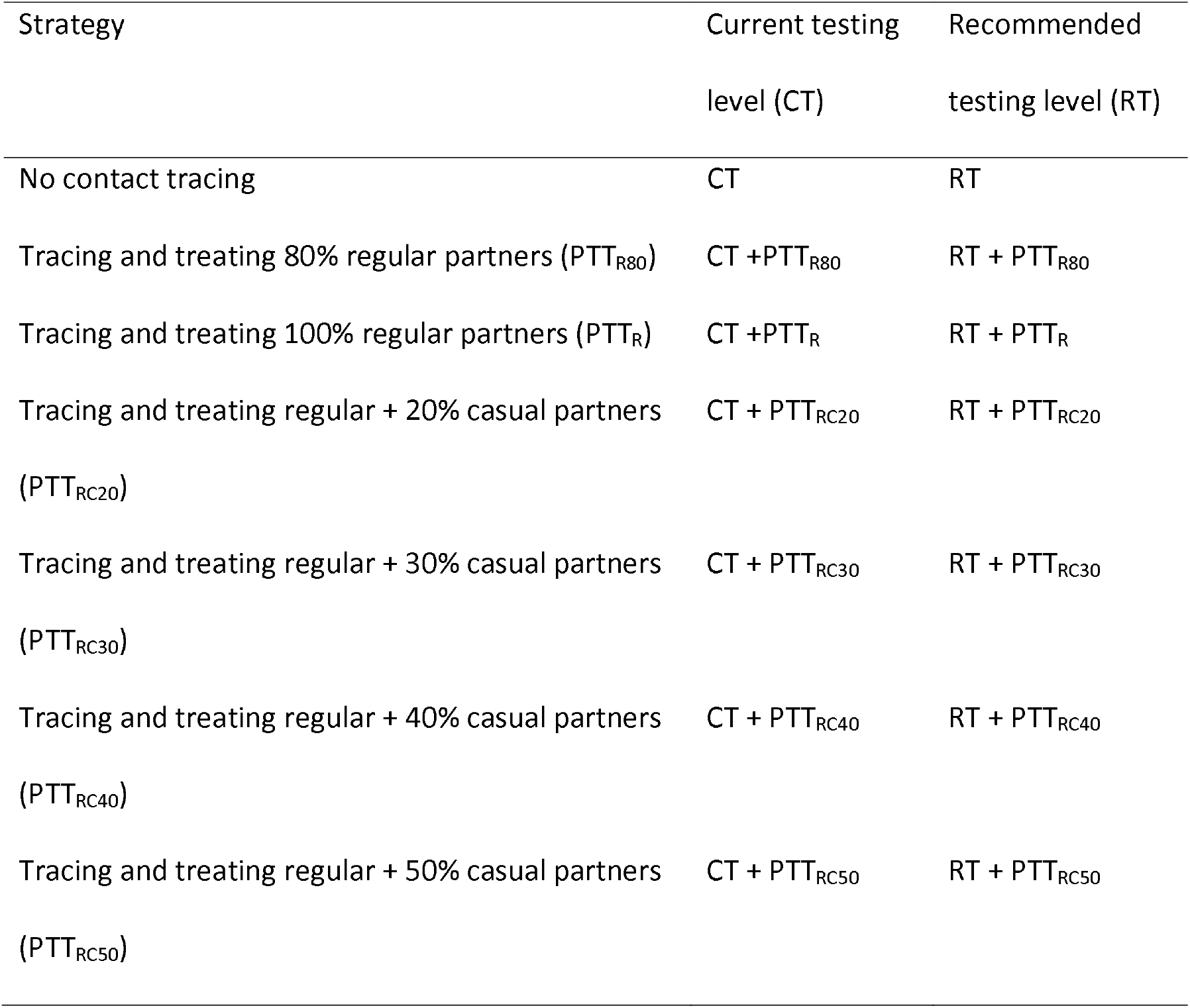
Outbreak response strategies and notation used

| Strategy | Current testing<br>level (CT) | Recommended<br>testing level (RT) |
| --- | --- | --- |
| No contact tracing | CT | RT |
| Tracing and treating 80% regular partners (PTT<sub>R80</sub>) | CT + PTT<sub>R80</sub> | RT + PTT<sub>R80</sub> |
| Tracing and treating 100% regular partners (PTT<sub>R</sub>) | CT + PTT<sub>R</sub> | RT + PTT<sub>R</sub> |
| Tracing and treating regular + 20% casual partners<br>(PTT<sub>RC20</sub>) | CT + PTT<sub>RC20</sub> | RT + PTT<sub>RC20</sub> |
| Tracing and treating regular + 30% casual partners<br>(PTT<sub>RC30</sub>) | CT + PTT<sub>RC30</sub> | RT + PTT<sub>RC30</sub> |
| Tracing and treating regular + 40% casual partners<br>(PTT<sub>RC40</sub>) | CT + PTT<sub>RC40</sub> | RT + PTT<sub>RC40</sub> |
| Tracing and treating regular + 50% casual partners<br>(PTT<sub>RC50</sub>) | CT + PTT<sub>RC50</sub> | RT + PTT<sub>RC50</sub> |

#### Outbreak Containment

The impact of outbreak response strategies on the ability of the imported strain to persist in the population are summarised in Table 3. Without enhanced public health measures (CT), the imported strain became extinct in 66% of simulations at 6 months post-importation, with a further 26% identified but persisting and 8% of outbreaks yet to be detected. By 2 years post-importation, 16% of simulated outbreaks still persisted, with mean population prevalence of 2.3% (IQR [0.5%-3.6%]). Almost 90% of this subset persisted at 5 years post-importation, reflecting endemic establishment (mean prevalence 14.7% (IQR [14.3%-16.7%]). Prevalence values for persisting simulations under the scenarios and time-points shown in Table 3 are reported in Table A12 of the Technical Appendix.

**Table 3.**
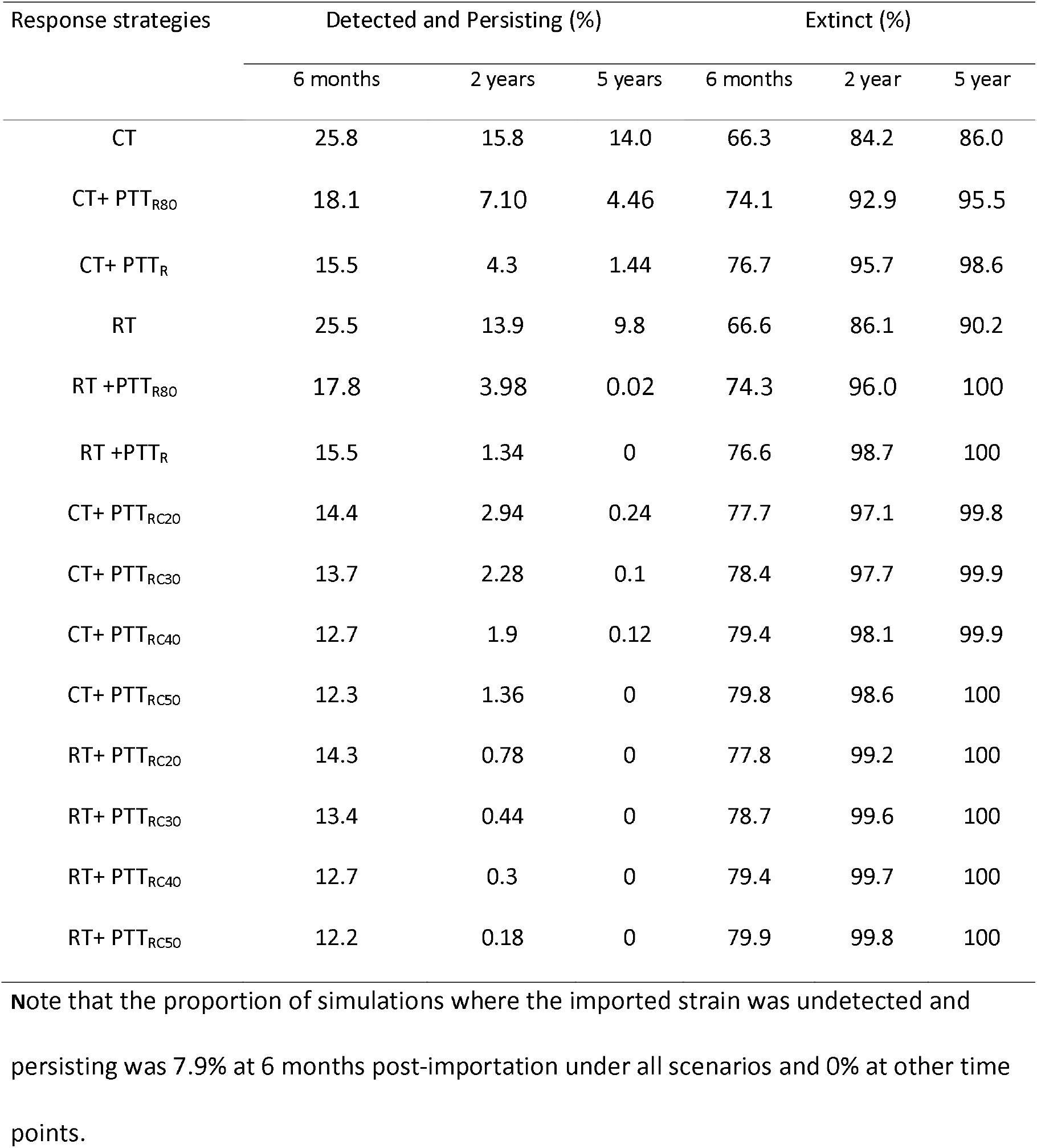
Proportion of simulations in which the imported strain was detected and persisting, or extinct at 6 months, 2 years and 5 years post-importation

All enhanced public health interventions increased the potential for control of the imported strain. Increasing testing to the recommended level (RT) had an increasing impact with time, with elimination of the imported strain rising from 84% to 86% and from 86% to 90% of simulations, at 2 years and 5 years respectively. This strategy, however, greatly reduced the size of the remaining outbreaks, with mean prevalence of 0.57% (IQR [0.1%-0.84%]) and 1.9% (IQR [0.8%-2.9%]) at 2 and 5 years, respectively, for the subset of simulations in which the imported strain persists. When current testing was supplemented by testing and treating 100% of regular partners (CT+PTTR), the probability of eliminating the imported strain increased to 96% and 98.5% of simulations and the mean prevalence of persisting strains was reduced to 0.25% (IQR [0.04%-0.38%]) and 0.45% (IQR [0.07%-0.78%]) at 2 and 5 years, respectively. The combination of recommended testing with testing/treating 100% of regular partners (RT+PTTR) led to elimination of the imported strain in 98.7% of simulations at 2 years, rising to 99.8% when supplemented by testing and treating 50% of casual partners in the last 2 months (RT+PTTRC50). In both these scenarios, the imported strain was eliminated in all 5000 simulations at 5 years post-importation.

Figure 3 provides a more detailed picture of detection and persistence over time for the interventions listed in Table 3. Panel A shows that the imported strain is either eliminated or are detected within 12 months of importation in all simulations. Under all eight control strategies (panels B and C), the proportion of simulations in which outbreaks have been detected but persist peaks at around 6 months post-importation. Inclusion of testing and treatment of regular partners greatly reduces the proportion of simulations in which the imported strain persists at each time point and this effect if further enhanced by testing/treatment of casual partners. The RT strategies have a modest impact alone but bring forward elimination of outbreaks when combined with partner treatment.

**Figure 3.**
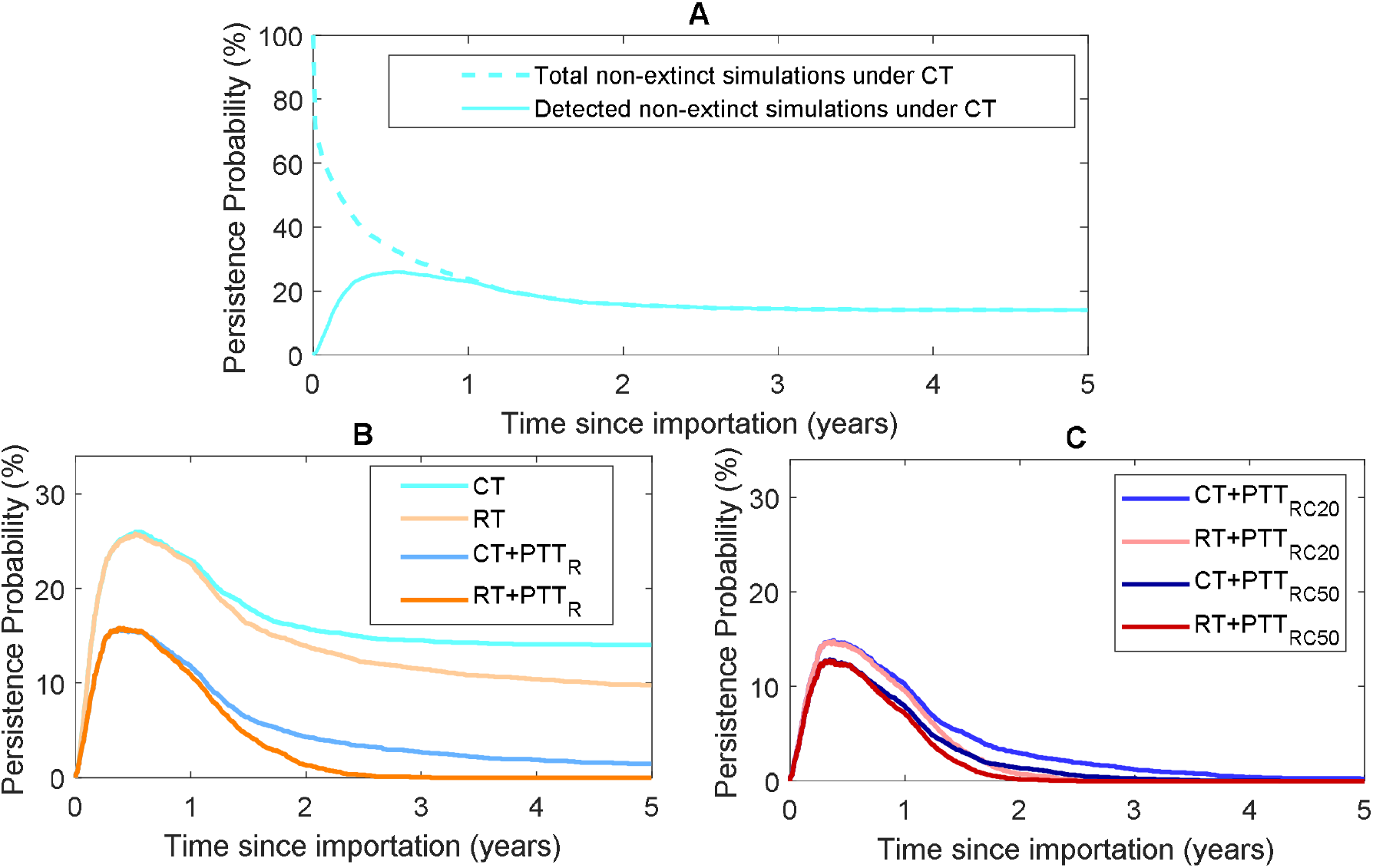
Panel A shows the persistence probability as a function of time under current testing. Dashed line: all simulations in which the imported strain persists, i.e., detected and undetected (100% at time = 0). Solid line: simulations in which the imported strain is detected and persisting (0% at time = 0). Dashed and solid lines converge at the end of the first year post-importation. Panel B shows trajectories for current and recommended testing with and without testing of regular partners. Panel C shows trajectories for current and recommended testing with testing of regular partners and a proportion of casual partners.

#### Outbreak duration and containment

In Figure 4 we show how success in eliminating the imported strain rises in quarterly periods after detection (panels A and B). Testing/treatment of regular partners is the most effective single strategy, increasing the probability of elimination within 3 months from 63% to 78% under CT when combined with partner treatment. This rises to 82% with testing/treatment of 50% of casual partners. To achieve >90% elimination at 12 months post-detection, RT in combination with testing/treatment of regular partners is required at a minimum. The probability of achieving elimination within 24 months of detection approaches 100% under strategies that additionally incorporate casual partner testing/treatment.

**Figure 4.**
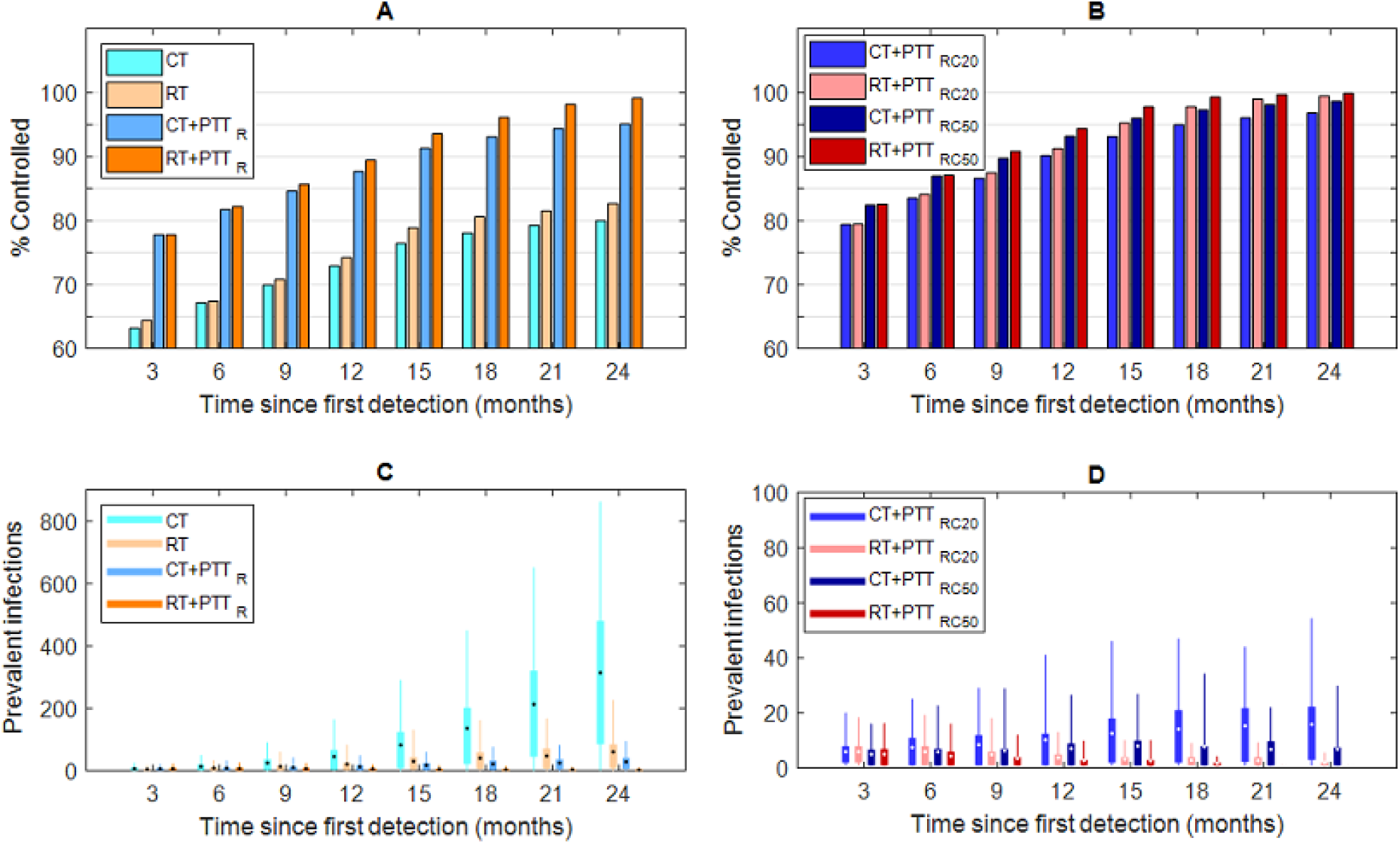
Panel A and B show the proportion of those simulations in which the imported strain is extinct, as a function of the time from the first detection and outbreak response strategy. Panel C and D shows the outbreak size of imported NG strain among these simulations as a function of time from the first detection and outbreak response strategy. The box denotes the interquartile range (25% to 75%), the whiskers the quantile (5% to 95%), the horizontal line in each box the median, and the dot in each box the mean. Outbreak size is the number or infected individuals at each time point for simulations in which the imported strain persists.

Panels C and D show how successful the various control strategies are in reducing the number of prevalent infections (outbreak size) at quarterly intervals. RT and/or addition of partner testing/treatment greatly reduce the growth in outbreak size in comparison to CT. Addition of RT to partner testing is more effective in constraining outbreak size than expanding testing and treatment to 20% or 50% of casual partners.

#### Sensitivity analysis

Sensitivity analysis is summarised in detail in section 3.2 of the technical appendix, covering the impact of less favourable assumptions regarding diagnosis and treatment, the impact of increased condom-use and the effect of a 6-month delay in public health response. Also, less favourable assumptions for diagnosis and treatment led to decreased effects of the interventions (Table A11). Increased condom-use had little impact with base-case parameters due to the strong role of oropharyngeal transmission but had greater effect with altered assumptions around symptomatic proportions (Table A13). Finally, all interventions were much less effective if implemented at a 6-month delay post-importation (Table A10).

## Discussion

In this modelling study we show that dissemination of emergent or imported strains of NG can be contained in a simulated Australian urban MSM population using moderately intense combinations of population-based and case-based testing/treatment strategies. The work is motivated by recent reports from Australia and the UK of imported XDR NG strains (5, 7, 16), including those with documented ceftriaxone/azithromycin dual treatment failure for oropharyngeal infection (7, 8) and the potential for outbreaks that are difficult and expensive to control (19). For instance, analysis of the historical rise in Australia of ciprofloxacin resistance (13) suggests that once resistant NG had become established overseas, variants of these resistant strains were repeatedly imported into Australia, leading eventually to a requirement to change the recommended treatment.

Under current testing practice, we find that around 1 in 7 imported NG strains will persist in the simulated MSM population at 5 years post-importation, with a mean prevalence of 15% at this point. However, both the probability of persistence and resulting prevalence can be greatly reduced by enhanced public health measures. When regular partners of NG cases are tested, as recommended in Australian contact tracing guidelines (27), the persistence probability drops to 0.5% at 5 years post-importation and falls further to 0.27% and 0%, respectively, if 20% or 50% of casual partners within the last 2 months can be tested/treated. These more intense case-based strategies are effective but need to be sustained for up to 2 years post-detection to eliminate >95% of imported strains. In the recent high-level azithromycin resistance NG outbreak in England (28), 118 total cases across both heterosexuals and MSM were investigated in an outbreak lasting more than 4 years. In this instance, ∼35% of partners of heterosexuals but none of MSM were recorded as being tested, suggesting that reaching the simulated testing levels evaluated here may be challenging.

A complementary approach is to increase the rate of background STI testing. Current testing rates in Australia are high when compared with other international settings (29), but are substantially lower than recommended in the Australian STI Management (24) and STIGMA (25) guidelines. Our results show that this approach in isolation is more effective at constraining the size of outbreaks than eliminating them. In Australia, it may be plausible to achieve the recommended testing level given that in 2018 around 90% of surveyed Sydney MSM undertook an STI test at least once per year (20), and that a large proportion of high-risk MSM access three-monthly STI testing in conjunction with the uptake of HIV pre-exposure prophylaxis (30). In addition, the new Australian STIGMA guidelines recommend quarterly testing for men with ∼4+ partners per year (25), which would greatly increase the proportion of the MSM population undertaking three-monthly STI testing.

Combining case-based strategies with increased population testing was beneficial for control, with all such simulations in which greater than 20% of casual partners were tested/treated, resulting in elimination of the imported strain within 5 years. If 50% of casual partners are tested/treated (RT+PTTRC50), elimination of the imported strain occurred within 2 years of detection in all simulations with no outbreak exceeding 20 prevalent infections during this period. These results highlight the importance of combined approaches using both targeted (partner testing and treatment) and population level strategies (increased community testing) to facilitate elimination. While not investigated here, synergistic effects between these interventions seem possible, with community testing reducing outbreak size and facilitating more intensive contact tracing.

In sensitivity analysis we also considered the effect of increasing condom use. This did not have a significant impact on control of the imported strain in the base case. This is because under the base case assumption that 90% of urethral cases are symptomatic and rapidly treated, urethral infection is relatively unimportant in overall transmission. When the fraction of urethral infections assumed to be symptomatic was reduced to 60%, urethral transmission became much more important and consequently condom use was found to be more effective.

The findings of this study should be interpreted in the context of the following limitations. The baseline (pre-intervention) model was calibrated to anatomical site-specific gonorrhoea prevalence as estimated by Zhang *et al*. (18), leading to inferred transmission probabilities, which qualitatively appear plausible but for which no quantitative estimates are available. These prevalence targets were chosen over incidence estimates from the ACCESS study (22) and were similar to more data-driven estimates of prevalence from Victoria (31) but we note that model produces similar site-specific incidence to 2018 ACCESS data.

In regard to diagnostic sensitivity and treatment success, we compared our base-case results with more pessimistic assumptions, which resulted in higher persistence probabilities (except for RT and RTSTIGMA at 5 years post initial importation), but typically slower outbreak growth. This occurred because as the same prevalence calibration targets were used for these simulations, assuming less effective treatment requires compensatory reductions in transmission probabilities compared to that of the best-case. Co-circulation of an endemic strain with the imported strain and any resultant interactions were not considered in this work, which focused on a single importation event. We note that there is limited evidence of mixed infections occurring at a single anatomical site but if this occurs it appears to be a rare occurrence (32). Sexual behaviour is fixed in time and assumed to be age-independent in this model. As the simulation timeframe is just 5 years, in comparison to a sexual lifetime of ∼50 years, we expect little impact of the former. In relation to age variation, this is likely implicitly captured by partner-change rates but could also influence testing rates and the risk of overseas acquisition, which are not considered here. In the base case, we assume that the public health strategies are implemented immediately upon detection of an imported strain. When implementation was delayed by 6 months, partner therapy and increased community testing were much less effective. Finally, we ignore bridging between MSM and heterosexual populations which could increase the likelihood of an imported strain entering the MSM population.

This is the first detailed exploration of the potential impacts of outbreak control strategies on the propagation of an imported NG strain. It suggests that a combination of increased case-based STI testing as well more regular background testing would be successful in eliminating onward spread from a very high proportion of importations. Australian guidelines for management of gonococcal infections include a heightened response to cases of ceftriaxone and/or high-level azithromycin resistance (33). This study provides additional evidence of the effect of these approaches alone and in combination with increases in background testing of asymptomatic MSM. Such increases have occurred in the context of the recent roll-out of pre-exposure prophylaxis for HIV in Australia (34) but our results do not support this being an effective strategy in isolation. However, both feasibility and resource utilisation need further investigation, in particular the capacity to deal with multiple and potentially concurrent importation events, such as occurred in the emergence of ciprofloxacin resistance in Australia during the 1990s.

## Methods

An overview of the modelling methodology is provided here, with complete details provided in the Technical Appendix.

### Gonorrhoea Natural History and Transmission

The natural history of gonorrhoea is captured in a Susceptible-Exposed-Infectious-Recovered (SEIRS) framework. Individuals enter the population in the susceptible ‘state’, and can move progressively through the exposed (infected but not yet infectious) and infectious states following sexual contact with an infected person, before entering the recovered state (immune to reinfection) following resolution of infection, and then finally returning to the susceptible state. We assume a brief duration of immunity reflecting evidence of weak immunity following infection (35) but frequent reinfection (36). Parameter values relating to gonorrhoea natural history have been derived, where possible, from published literature and are listed in Table 1.

Transmission is assumed to be possible between each pairing of anatomical sites leading to eight possible routes of transmission (See Table A5, Technical Appendix). Infections at different anatomical sites within a given individual are assumed to be localised and independent, such that infection at one anatomical site does not influence the properties of infection (e.g., duration of infection) at any other anatomical site.

### Population and Sexual Behaviour

The model simulates a dynamic network of individuals connected via sexual partnerships, where both long-and short-term partnerships are considered, designated as ‘regular’ and ‘casual’, respectively. Individuals enter the sexually active population at age 16 years and on reaching age 65 years are replaced with a new individual aged 16 years, having the same sexual behaviour profile as the replaced individual.

On entry to the population, individuals are assigned a partner-type preference. Preference can be for regular partners only, casual partners only, or both types of partner. Where relevant, individuals are assigned a casual partner acquisition rate (CPAR). Individuals are also assigned a positional preference for anal sex (receptive/insertive/no preference). Partnership durations for each partnership type are assigned at formation by sampling from a distribution (see Table A2, Technical Appendix). Partner preferences and the CPAR distribution were based on data reported in the Gay Community Periodic Survey (GCPS) Sydney 2018 (20) and the Health In Men (HIM) Study (21), respectively. Group sex (37) is included as a subclass of casual partnership that only lasts for one day. Except for group sex, individuals can have at most one regular partner and/or one casual partner concurrently. An individual’s sexual behaviour remains constant over time.

Within partnerships, individuals can engage in a variety of sexual acts (e.g., anal sex, oral-genital and oral-anal sex, kissing) that facilitate transmission. The frequencies of these acts during partnerships are based on data reported by Phang *et al*. (38) and Rosenberger *et al*. (39). Kissing is an important transmission routine for oropharyngeal infection (40, 41), and if it is not included, the model is not able to catch the high prevalence of oropharyngeal infection.

In the baseline model, individuals are tested for gonorrhoea, at a rate based on data reported in GCPS Sydney 2018 (20) and by the Australian Collaboration for Coordinated Enhanced Sentinel Surveillance of Sexually Transmissible Infections and Blood-borne Viruses (ACCESS) (23). Condom use by partnership type is also derived from data reported in GCPS Sydney 2018 (20).

### Model Calibration

The model was calibrated to estimated anatomical site-specific NG prevalence in a hypothetical community sample as reported in Zhang *et al*. (18): oropharynx 8.6%; anorectum 8.3%; urethra 0.26%. This involved generating 10,000 parameter sets sampled from pre-specified ranges (Table A9, Technical Appendix) and then determining the set with the smallest mean-squared difference between model-generated prevalence and the target values. This calibration establishes the baseline model representing the current situation, where gonorrhoea is endemic in the population, prior to the importation/emergence of a new strain. While the motivation for this work is the threat of new strains that are resistant to current first-line treatments, we are interested here in assessing the effectiveness of outbreak response strategies, assuming imported or emergent strains are still treatable with second-line drugs such as carbapenems (7).

### Simulation Process

Results for each intervention scenario consist of 5,000 model simulations. Initially, each simulation is run for 10 years to establish the partnership network. After introduction of the imported strain, simulations are run with current testing rates until first detection of the imported strain, at which point the desired intervention is initiated and the simulation run for a further 5 years. Note that the imported strain can be detected when symptomatic patients seek for treatment and asymptomatic patients take regular test.

At any time-point, a simulation is categorised as being in one of four states regarding the status of the imported strain: 1) *undetected and persisting*; 2) *undetected and extinct*; 3) *detected and persisting*; and 4) *detected and extinct*. All simulations are initiated in the *undetected* and *persisting* state, with transition to *detected and persisting* occurring at the first positive test and transition to *extinct* (*detected or undetected*) when the last case resolves.

## Data Availability

The authors confirm that the data supporting the findings of this study are available within the article its supplementary material.

## References

1. Unemo M, Shafer WM. Antimicrobial resistance in Neisseria gonorrhoeae in the 21st century: past, evolution, and future. Clin Microbiol Rev. 2014;27(3):587–613.

2. Centers for Disease Control and Prevention. Antibiotic Resistance Threats in the United States 2013 [Available from: https://www.cdc.gov/drugresistance/threat-report-2013/index.html.

3. World Health Organization. Global action plan to control the spread and impact of antimicrobial resistance in Neisseria gonorrhoeae 2013 [Available from: http://www.who.int/reproductivehealth/publications/rtis/9789241503501/en/.

4. Lahra MM, Martin I, Demczuk W, Jennison AV, Lee KI, Nakayama SI, et al. Cooperative recognition of internationally disseminated ceftriaxone-resistant Neisseria gonorrhoeae strain. Emerg Infect Dis. 2018;24(4):735–40.

5. Whiley DM, Jennison A, Pearson J, Lahra MM. Genetic characterisation of Neiserria gonorrhoeae resistant to both ceftriaxone and azithromycin, Australia 2018. Lancet Infect Dis. 2018;18(7):717–8.

6. Jennison AV, Whiley D, Lahra MM, Graham RM, Cole MJ, Hughes G, et al. Genetic relatedness of ceftriaxone-resistant and high-level azithromycin resistant Neisseria gonorrhoeae cases, United Kingdom and Australia, February to April 2018. Eurosurveillance. 2019;24(8):1900118.

7. Eyre DW, Town K, Street T, Barker L, Sanderson N, Cole MJ, et al. Detection in the United Kingdom of the Neisseria gonorrhoeae FC428 clone, with ceftriaxone resistance and intermediate resistance to azithromycin, October to December 2018. Euro Surveill. 2019;24(10).

8. Poncin T, Fouere S, Braille A, Camelena F, Agsous M, Bebear C, et al. Multidrug-resistant Neisseria gonorrhoeae failing treatment with ceftriaxone and doxycycline in France, November 2017. Euro Surveill. 2018;23(21).

9. Rob F, Klubalova B, Nycova E, Hercogova J, Unemo M. Gentamicin 240 mg plus azithromycin 2 g vs. ceftriaxone 500 mg plus azithromycin 2 g for treatment of rectal and pharyngeal gonorrhoea: a randomized controlled trial. Clin Microbiol Infect. 2020;26(2):207–12.

10. Lewis DA. New treatment options for Neisseria gonorrhoeae in the era of emerging antimicrobial resistance. Sexual Health. 2019;16(5):449–56.

11. Hook EW, III, Golden MR, Taylor SN, Henry E, Tseng C, Workowski KA, et al. Efficacy and Safety of Single-Dose Oral Delafloxacin Compared With Intramuscular Ceftriaxone for Uncomplicated Gonorrhea Treatment: An Open-Label, Noninferiority, Phase 3, Multicenter, Randomized Study. Sex Transm Dis. 2019;46(5):279–86.

12. Lahra MM, Ward A, Trembizki E, Hermanson J, Clements E, Lawrence A, et al. Treatment guidelines after an outbreak of azithromycin-resistant Neisseria gonorrhoeae in South Australia. Lancet Infect Dis. 2017;17(2):133–4.

13. Hanrahan JK, Hogan TR, Buckley C, Trembizki E, Mitchell H, Lau CL, et al. Emergence and spread of ciprofloxacin-resistant Neisseria gonorrhoeae in New South Wales, Australia: lessons from history. J Antimicrob Chemother. 2019;74(8):2214–9.

14. Moodley P, Sturm AW. Ciprofloxacin-resistant gonorrhoea in South Africa. Lancet. 2005;366(9492):1159.

15. Tapsall JW, Limnios EA, Murphy D. Analysis of trends in antimicrobial resistance in Neisseria gonorrhoeae isolated in Australia, 1997-2006. J Antimicrob Chemother. 2008;61(1):150–5.

16. Horner P, Fifer H, Nathan B, Eyre D, Town K, Mohammed H, et al. P675 Two recent cases of extensively drug-resistant (XDR) gonorrhoea in the united kingdom linked to a european party destination. Sex Transm Infect. 2019;95(Suppl 1):A295–A6.

17. Hui BB, Fairley CK, Chen MY, Grulich AE, Hocking JS, Prestage GP, et al. Oral and anal sex are key to sustaining gonorrhoea at endemic levels in MSM populations: a mathematical model. Sex Transm Infect. 2015;91(5):365–9.

18. Zhang L, Regan DG, Chow EPF, Gambhir M, Cornelisse V, Grulich A, et al. Neisseria gonorrhoeae transmission among men who have sex with men: an anatomical site-specific mathematical model evaluating the potential preventive impact of mouthwash. Sex Transm Dis. 2017;44(10):586–92.

19. Regan DG, Hui BB, Wood JG, Fifer H, Lahra MM, Whiley DM. Treatment for pharyngeal gonorrhoea under threat. Lancet Infect Dis. 2018;18(11):1175–7.

20. Broady T, Mao L, Lee E, Bavinton B, Keen P, Bambridge C, et al. Gay Community Periodic Survey: Sydney 2018. Centre for Social Research in Health, UNSW Sydney; 2018.

21. Fogarty A, Mao L, Zablotska I, Santana HR, Prestage G, Rule J, et al. The Health in Men and Positive Health cohorts: a comparison of trends in the health and sexual behaviour of HIV-negative and HIV-positive gay men, 2002-2005. National Centre in HIV Social Research, UNSW Sydney; 2006.

22. Callander D, Guy R, Fairley CK, McManus H, Prestage G, Chow EPF, et al. Gonorrhoea gone wild: rising incidence of gonorrhoea and associated risk factors among gay and bisexual men attending Australian sexual health clinics. Sex Health. 2019;16(5):457–63.

23. Callander D, Donovan B, Guy R. The Australian Collaboration for Coordinated Enhanced Sentinel Surveillance of Sexually Transmissible Infections and Blood Borne Viruses: NSW STI report 2007-2014. Sydney, NSW, Australia: Kirby Institute, UNSW Sydney; 2015.

24. Australasian Sexual Health Alliance. Australian STI Management Guidelines 2018 [Available from: http://www.sti.guidelines.org.au/.

25. STIGMA. Australian STI & HIV Testing Guidelines 2019 for Asymptomatic MSM 2019 [Available from: https://stipu.nsw.gov.au/wp-content/uploads/STIGMA_Guidelines2019_Final-1.pdf.

26. Australian Contact Tracing Guidelines [Available from: http://www.contacttracing.ashm.org.au/.

27. Hui B, Fairley C, Chen M, Grulich A, Hocking J, Prestage G, et al. Oral and anal sex are key to sustaining gonorrhoea at endemic levels in MSM populations: a mathematical model. Sex Transm Infect. 2015;91(5):365–9.

28. Smolarchuk C, Wensley A, Padfield S, Fifer H, Lee A, Hughes G. Persistence of an outbreak of gonorrhoea with high-level resistance to azithromycin in England, November 2014lllMay 2018. 2018;23(23):1800287.

29. World Health Organization (WHO). Guidelines: Prevention and Treatment of HIV and Other Sexually Transmitted Infections Among Men Who Have Sex with Men and Transgender People: Recommendations for a Public Health Approach 2011. 2011 [Available from: https://www.ncbi.nlm.nih.gov/books/NBK304456/.

30. The Australasian Society of HIV, Viral Hepatitis and Sexual Health Medicine, (ASHM). PrEP Guidelines Update. Prevent HIV by Prescribing PrEP. Sydney, 2019. Sydney [Available from: https://ashm.org.au/resources/hiv-resources-list/prep-guidelines-2019/.

31. Chow EPF, Tomnay J, Fehler G, Whiley D, Read TRH, Denham I, et al. Substantial increases in chlamydia and gonorrhea positivity unexplained by changes in individual-level sexual behaviors among men who have sex with men in an Australian sexual health service from 2007 to 2013. Sex Transm Dis. 2015;42(2):81–7.

32. Goire N, Kundu R, Trembizki E, Buckley C, Hogan TR, Lewis DA, et al. Mixed gonococcal infections in a high-risk population, Sydney, Australia 2015: implications for antimicrobial resistance surveillance? J Antimicrob Chemother. 2016;72(2):407–9.

33. Communicable Diseases Network Australia (CDNA). The Series of National Guidelines (SoNG): Gonococcal Infection (Appendix G) 2019 [Available from: https://www1.health.gov.au/internet/main/publishing.nsf/Content/cdnasongs.htm.

34. Department of Health, Australian Government,. Pharmaceutical Benefits Scheme (PBS) [Available from: http://www.pbs.gov.au/pbs/home.

35. Liu Y, Feinen B, Russell M. New Concepts in Immunity to Neisseria Gonorrhoeae: Innate Responses and Suppression of Adaptive Immunity Favor the Pathogen, Not the Host. Front Microbiol. 2011;2:52.

36. Lister NA, Smith A, Tabrizi SN, Garland S, Fairley CK. Re-infection of Neisseria gonorrhoeae and Chlamydia trachomatis infections among men who have sex with men. Int J STD AIDS. 2006;17(6):415–7.

37. Wilson DP, Prestage G, Donovan B, Gray RT, Hoare A, McCann PD, et al. Phase A of the National Gay Men’s Syphilis Action Plan: modelling evidence and research on acceptability of interventions for controlling syphilis in Australia. Sydney: National Centre in HIV Epidemiology and Clinical Research, UNSW Sydney; 2009.

38. Phang CW, Hocking J, Fairley CK, Bradshaw C, Hayes P, Chen MY. More than just anal sex: the potential for sexually transmitted infection transmission among men visiting sex-on-premises venues. Sex Transm Infect. 2008;84(3):217–9.

39. Rosenberger JG, Reece M, Schick V, Herbenick D, Novak DS, Van Der Pol B, et al. Sexual behaviors and situational characteristics of most recent male-partnered sexual event among gay and bisexually identified men in the United States. J Sex Med. 2011;8(11):3040–50.

40. Chow EP, Cornelisse VJ, Williamson DA, Priest D, Hocking JS, Bradshaw CS, et al. Kissing may be an important and neglected risk factor for oropharyngeal gonorrhoea: a cross-sectional study in men who have sex with men. Sex Transm Infect. 2019;95(7):516–21.

41. Fairley CK, Cornelisse VJ, Hocking JS, Chow EP. Models of gonorrhoea transmission from the mouth and saliva. The Lancet Infectious Diseases. 2019;19(10):e360–e6.

42. Hook EW, III., Handsfield HH. Gonococcal Infections in the Adult. In: Holmes KK, Sparling PF, Stamm WE, Piot P, Wasserheit JN, Corey L, et al., editors. Sex Transm Dis. New York: McGraw Hill; 2008.

43. Ryder N, Lockart IG, Bourne C. Is screening asymptomatic men who have sex with men for urethral gonorrhoea worthwhile? Sexual Health. 2010;7(1):90–1.

44. Rebe K, Lewis D, Myer L, de Swardt G, Struthers H, Kamkuemah M, et al. A Cross Sectional Analysis of Gonococcal and Chlamydial Infections among Men-Who-Have-Sex-with-Men in Cape Town, South Africa. PLoS One. 2015;10(9):e0138315.

45. Dudareva-Vizule S, Haar K, Sailer A, Wisplinghoff H, Wisplinghoff F, Marcus U, et al. Prevalence of pharyngeal and rectal Chlamydia trachomatis and Neisseria gonorrhoeae infections among men who have sex with men in Germany. Sex Transm Infect. 2014;90(1):46–51.

46. Fairley CK, Chen MY, Bradshaw CS, Tabrizi SN. Is it time to move to nucleic acid amplification tests screening for pharyngeal and rectal gonorrhoea in men who have sex with men to improve gonorrhoea control? Sexual Health. 2011;8(1):9–11.

47. Chow EP, Camilleri S, Ward C, Huffam S, Chen MY, Bradshaw CS, et al. Duration of gonorrhoea and chlamydia infection at the pharynx and rectum among men who have sex with men: a systematic review. Sex Health. 2016;13(3):199–204.

48. Fairley CK, Chow EP, Hocking JS. Early presentation of symptomatic individuals is critical in controlling sexually transmissible infections. Sexual Health. 2015;12(3):181–2.

49. Harrison WO, Hooper RR, Wiesner PJ, Campbell AF, Karney WW, Reynolds GH, et al. A trial of minocycline given after exposure to prevent gonorrhea. N Engl J Med. 1979;300(19):1074–8.

50. Mehta SD, Erbelding EJ, Zenilman JM, Rompalo AM. Gonorrhoea reinfection in heterosexual STD clinic attendees: longitudinal analysis of risks for first reinfection. Sex Transm Infect. 2003;79(2):124–8.

51. Hedges SR, Mayo MS, Mestecky J, Hook EW, 3rd, Russell MW. Limited local and systemic antibody responses to Neisseria gonorrhoeae during uncomplicated genital infections. Infect Immun. 1999;67(8):3937–46.

52. Cornelisse VJ, Chow EP, Huffam S, Fairley CK, Bissessor M, De Petra V, et al. Increased detection of pharyngeal and rectal gonorrhea in men who have sex with men after transition from culture to nucleic acid amplification testing. Sex Transm Dis. 2017;44(2):114–7.

