## Supplementary for "Modelling outbreak response strategies for preventing spread of emergent *Neisseria gonorrhoeae* strains in men who have sex with men in Australia"

Technical Appendix

### 1. Individual-based Model

#### 1.1 Overview

An individual-based, anatomical site-specific mathematical model was developed to simulate the transmission of gonorrhoea among Australian MSM. The model represents a dynamic network of sexually active individuals that are linked by sexual partnerships. The model consists of two main components, i.e., individuals and the sexual partnership network.

Each individual in the model is specified by the following features.

- Age (16 – 65 years old)
- Infection status (susceptible, exposed, infectious, recovered) by anatomical site (pharynx, urethra, rectum).
- Sexual behaviours, including preferred partnership type (regular only, casual only, both), casual partner acquisition rate per 6 months and anal sex role (insertive, receptive and versatile).
- Information about partnership formation and dissolution, including historical partnerships in last 6 months, start and end days of partnerships, availability for new partnerships and the probability of acquiring new partners.
- Intervention features, including testing frequency and condom usage.

Moreover, individuals are linked by sexual partnerships, which we specify as follows:

- Partnership type (casual or regular partnership).
- Partnership duration (duration distributions differ by partnership type).
- Scheduled sexual acts for every day during the partnership (kissing, oral sex, rimming, anal sex and docking).

The model tracks the partnership network, and the sexual activities and infection status of individuals on a daily basis. Figure 1 (in main text) shows how the model operates in each simulation cycle, while Algorithm 1 outlines the approach to modelling infection in time. For convenience of implementation and analysis, we set each month to be 30 days in length (one year is then 12 months or 360 days).

At the beginning of each simulation cycle, the status of each individual and the sexual partnership network are imported from the previous simulation step. For each partnership, sexual acts are scheduled and performed, and transmissions from infectious to susceptible individuals within partnerships are enumerated. These infection events lead to a change of infection status of the susceptible individual, with durations of infection sampled from the duration distribution for each newly infected site.

At the end of the simulation cycle, any events such as natural recovery, screening and treatment of infection or replacement of new individual are processed and the status of individuals are updated accordingly. Also, partnerships that were scheduled to separate on that day are separated, and individuals available for new partnerships are selected to form new partnerships. Once the sexual partnership network and properties of individuals have been updated, the model moves to next simulation cycle.

The Algorithm 1 shows the framework of the model.

**Algorithm 1. Individual-based Model**

1. **Initialization of individuals and sexual partnership network.**

At $day=0$, generate N (N=10,000) individuals with properties being randomly initialized and store individuals into available list $Ra$ and $Ca$ according to their availability of regular and casual partnership.

Randomly select and remove two individuals from available list $Ra$ and $Ca$, and form the preferred partnership. This process is repeated until <2 individual left on each list.

2. Equilibrium of sexual partnership

For $day=1:d_{network}$

a. Aging and replacement.

Check the age of each individual and remove the individual when its age reaches 65 and replace it with a new and uninfected individual aged 16. The new individual has the same properties as the one it replaces. And all the partnerships of the old individual cease.

b. Partnership separation and formation.

**Separation**. Break the regular and casual partnerships that had been scheduled to stop on the $day$ and put corresponding individuals into available list $Ra$ or $Ca$.

**Formation**. As described in the Algorithm 2 and 3.

End for

(Remark:$d_{network}=5 years\times360$. At the end of Step 2 the dynamic sexual partnership network is expected to reach the equilibrium states, which is shown in Figure 2.)

3. Introduction of background strains

Randomly introduce local infections to the population, where ~7%, 7% and 0.5% of individuals are allocated with pharyngeal, rectal and urethral infections, respectively, so that the transmission of local infection can reach the equilibrium quickly. Note that this allocation is not mutually exclusive, so that some individuals may be allocated infections at $\geq2$ sites.

4. Equilibrium of gonorrhoea transmission

For $day=\left( d_{network}+1 \right) :{(d}_{network}+d_{infection})$

a. Scheduling and performance of sex acts and infection transmission.

Go through every partnership and schedule and conduct sex acts. Allocate disease transmission events based on sexual acts and associated transmission probabilities.

b. Aging and replacement.
c. Partnerships separation and formation.
d. Recovery, screening and treatment.

For each infected individual, the asymptomatic infection is naturally cleared (recovery) or detected and treated when patients have regular STI tests. The symptomatic infections are treated when patients seek treatment, and infections at other sites are also treated

e. Daily prevalence calculation

End for

(Remark: $d_{infection}=5 years \times360.$ At the end of Step 4, gonorrhoea transmission is expected to reach dynamic equilibrium as shown in Figure 4)

5. Introduction of invading strain

Randomly select an infected individual according to Algorithm 4 and change the status of all other infected individuals to be susceptible. Run the model forward under various outbreak response strategies and compare their impact.

(Detailed process is showed in the Algorithm 3)

6. Evaluation of outbreak response strategies.

For $day={(d}_{network}+d_{infection}+1) :{(d}_{network}+d_{infection}+d_{intervention})$

a. Scheduling and performance of sex acts and infection transmission.
b. Aging and replacement
c. Partnership separation and formation
d. Recovery, screening and treatment
e. Output of outbreak information

End for

(Remark: $d_{intervention}=5\times360.$)

#### 1.2 Infection Dynamics

#### In the model we used an SEIRS (*Susceptible 🡪 Exposed🡪 Infectious 🡪 Recovered 🡪 Susceptible*) structure to describe the natural history of gonorrhoea. When a susceptible individual is infected with gonorrhoea, he experiences an *exposed* period, during which he is infected but not infectious, before becoming *infectious*. The individual may then experience either asymptomatic or symptomatic gonorrhoea, with symptomatic proportions varying by site of infection and with the duration of infection assumed to be independent of symptom status but varying by site with the rectal infection assumed to have longer duration. Asymptomatic gonorrhoea can be spontaneously cleared or treated after taking a test. For symptomatic gonorrhoea, we assume an *incubation* period that includes the *exposed* period and a small fraction of the *infectious* period in the model dynamics as once symptoms occur, the individual is assumed to take a test and then be treated within a short period. We assume a short period of immunity following infection before returning to susceptible status. Finally, we assume that any time and individual is tested for gonorrhoea this is applied to all anatomical sites, with treatment applied if relevant. Parameters governing gonorrhoea infection are summarized in Table A1.

**Table A1 Parameters of gonorrhoea infection**

| Parameter description | Value (bounds) | Source/Assumption |
| --- | --- | --- |
| *Symptomatic proportion* | | |
| Pharyngeal infection | 0% | Pharyngeal gonorrhoea infection has mild and non-specific symptoms. |
| Urethral infection | 90% | [1, 2]. |
| Rectum infection | 12% | [3] |
| *Durations (days)* | | |
| Pharyngeal infection | 84 (70,138) | Assume to follow Gamma (201,0.4) within bounds based on [4, 5]. |
| Urethral infection | 84 (70, 140) | Assume to follow Gamma (206,0.4) within bounds based on model assumption [6, 7], |
| Rectal infection | 343 (336,361) | Assume to follow Gamma (3702,0.1) within bounds based on [4], |
| Treatment of symptomatic infection | 3 (1,7) | [8], and assume to follow Gamma (3, 0.86) within bounds. The parameter is the period that patients seek treatment for symptomatic urethral or rectal infections. |
| Incubation | 4 (2,10) | Assume to follow Exp(4) within bounds based on [9]. Note that the exposed period is shorter than the incubation period. |
| Exposed | 3.6 (1,9) | Assume to be uniformly distributed from 1 to incubation period. |
| Immunity | 3.5 (1,7) | Assume to be uniformly distributed within bounds based on recommended period of abstinence post treatment [10]. |

#### 1.3 Sexual Partnership and Population

There are two types of sexual partnerships considered in our model: regular (long-term) partnerships and casual (short-term) partnerships. Group sex is also considered and implemented as a specific type of casual partnership.

Individuals are allowed to have at most one regular partner and/or at most one casual partner at the same time, except in group sex, where they can have multiple casual partners simultaneously. Casual partnerships that involve group sex have a duration of only one day. We assume that the sexual behaviour (partnership preference, partner acquisition rate and anal sex preference) of individuals is constant in time and is not affected by the characteristics of their partners. In real populations, of course individual preferences do change over time. However, as the timeframe of gonorrhoea infection is relatively short, we do not expect strong effects of individual changes in preference on dynamics. We also note that these preferences are informed by population level data sources, not at an individual level. Information about sexual partnerships are listed in Table A2.

**Table A2 Parameters of MSM sexual partnerships**

| Parameter | Value | Source/Assumption |
| --- | --- | --- |
| *Regular partnership (RP)* | | |
| Population size | 62.6% of all MSM | Assume based on Gay Community Periodic Survey (GCPS) 2018 Sydney [11], where **26.8%** have only regular partners and **35.8%** have both types of partners |
| Duration | 4 years (=1440 days) | Assume to be follow $Exp\left( \frac{1}{1440} \right)$ based on Hui et al. 2015 [6] and [12] |
| *Casual partnership (CP)* | | |
| Population Size | 59.3% of all MSM | Assume based on GCPS 2018 Sydney [11], where **23.5%** have only casual partners and **35.8%** have both types of partners |
| Casual Partner Acquisition Rate per 6 months (*CPAR*) | See Figure A1 | From Wilson et.al 2009 [13], which was adapted from Fogarty et al. 2006 [14]. |
| Duration | 1-14 days | Assume based on individual’s acquisition rate per 6 months but the longest duration is less than 14 days. |
| *Group sex* | | |
| Population Size | ~31% of all MSM | Assume based on Wilson et al. [13], where group sex is a type of casual partnership, and individuals who have 5+ casual partners per 6 months are assumed to involve in group sex. This assumption result in ~31% of MSM engaged in group sex. |
| Group size | 4.4 | Assume to follow generalized Pareto distribution based on Wilson et al. [13]. |
| Duration | 1 day | Assume the group sex partnership only last for 1 day. |
| *Single population* | | |
| Population Size | 13.9% of all MSM | Assume based on GCPS 2018 Sydney [11], where 13.9% of MSM have no sex partners in last 6 months. |
| CPAR | <0.5 | Assume that single population can be involved in casual partnership but with a small acquisition rate less than 0.5 per 6 months. |

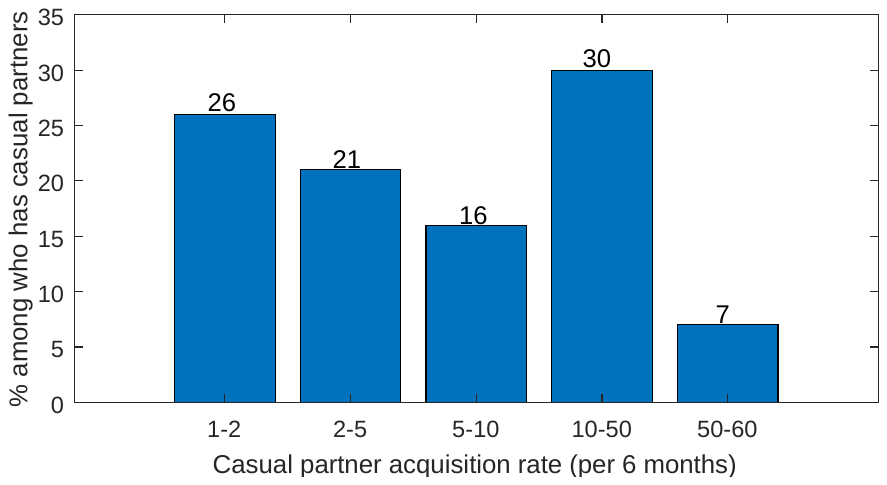

Figure A1. Casual Partner Acquisition Rate per 6 months (CPAR). The values are obtained from Wilson et.al 2009 [13], which was originally adapted from Fogarty et al. 2006 [14].

##### Regular Partnership

The list $Ra$ in Algorithm 1 is used to store all the individuals available for regular partnership. At each time point, two individuals are randomly selected from the list, if they were not partners of each other in the last partnership, they would form a new regular partnership. The duration of the regular partnership is randomly generated from an exponential distribution with mean of 1440 days. For example, when individuals $i$ and $j$ are to form a new regular partnership, the duration $dr_{i,j}$ of this partnership is as follows

$$dr_{i,j}=\left\lceil-log\left( U \right)\times1440 \right\rceil, U\sim U\left[ 0,1 \right].$$

After this period, the regular partnership would be separated, and the related individuals $i$ and $j$ are put back to the available list $Ra$. The formation of regular partnership is shown in Algorithm 2.

**Algorithm 2. Formation of regular partnerships**

1. Set$Ra_{d}=R_{a}$.
2. While ( $\left| Ra_{d} \right|^{*}>1$)

Randomly select two individuals from list $Ra_{d}$;

If (two individuals were not partners of each other in the last partnership)

Form a regular partnership for these two individuals;

Generate the duration of regular partnership;

End if

Remove these two individuals from$Ra_{d}.$

End while

3. Set$R_{a}=Ra_{d}$.

^*^ $\left| \cdot\right|$ denotes the number of elements in a list.

##### Casual Partnership

The formation of casual partnership depends on CPAR from Wilson et.al 2009 [13], as shown in Table A2.1. In the initialization step of Algorithm 1, each individual who can have casual partners would be assigned a CPAR as a feature of sexual behaviour. Note that in the model we categorize the population into four groups in terms of partnership preference (regular only, casual only, both and single), and individuals from three groups (casual only, both, and single) can have casual partners. The assignment of CPAR is as shown in Algorithm 3.

**Algorithm 3. Assignment of CPAR**

For $i=1:N$

If (individual $i$ is from the group of having casual only or both)

Generate a random number $X_{i}$ from uniform distribution $U\left[ 0,1 \right].$

If ${(0<X}_{i}\leq0.26)$

$${CPAR}_{i}=1+U, U\sim U\left[ 0,1 \right],$$

Else if ${(0.26<X}_{i}\leq0.47$)

$${CPAR}_{i}=2+3\times U, U\sim U\left[ 0,1 \right],$$

Else if ${(0.47<X}_{i}\leq0.63$)

$${CPAR}_{i}=5+5\times U, U\sim U\left[ 0,1 \right],$$

Else if ${(0.63<X}_{i}\leq0.93$)

$${CPAR}_{i}=10+40\times U, U\sim U\left[ 0,1 \right],$$

Else if ${(0.93<X}_{i}\leq1$)

$${CPAR}_{i}=50+10\times U, U\sim U\left[ 0,1 \right],$$

End if

Else if (individual $i$ is from the group of single)

$${CPAR}_{i}=0.5\times U, U\sim U\left[ 0,1 \right],$$

Else

$${CPAR}_{i}=0.$$

End if

End for

In Algorithm 1, each individual available for casual partnership in $Ca$ has a probability of forming a new casual partnership (and group sex) each day. The probability of individual $i$ getting involved in a casual partnership is defined as follows and is updated for each day$d$.

$${Pc}_{i}=\max\left\{ \frac{CPAR-NCP}{180-\left( d -Lday_{180} \right)}, 0 \right\},$$

where $NCP$ is total number of casual partners in last 180 days (from day $d-179$ to day $d$) and $Lday_{180}$ is the last day of the first casual partnership in last 180 days. If the total number of casual partners in last 180 days is larger than acquisition rate, then ${Pc}_{i}$ is set to 0. Note that $NCP$ includes casual partners acquired from group sex. If the *CPAR* is less than 0.5, which characterises individuals that are single more than half the time each year in the population, the probability is defined differently as follows.

$${Pc}_{i}=\frac{1}{\frac{180}{CPAR}-\left( d-Lday \right)},$$

where $Lday$ is the day on which the individual’s last casual partnership ended.

Once${Pc}_{i}=1$, individual $i$, regardless of its *CPAR*, would get a casual partner with the priority in the simulation cycle. If individual $i$ fails to get a casual partner, ${Pc}_{i}$ would not change and remain at 1 for the next simulations cycle.

The durations between casual partnerships are also determined by *CPAR*. For example, an individual has 2 casual partners per 6 months, then the average time interval between two casual partnerships would be around 90 days. We assume that only after such period would an individual seek next casual partner. This time interval is randomly generated for individual $i$ according to partner acquisition rates as follows

$${dp}_{i}=\left\lceil-log\left( U \right)\times\frac{180}{{CPAR}_{i}} \right\rceil, U\sim U\left[ 0,1 \right].$$

Note that the interval would be large if the *CPAR* is small.

These intervals are then used to construct the duration of casual partnerships, where the duration of a casual partnership between individuals $i$ and $j$ defined as follows

$$dp_{ij}=min\left( {dp}_{i},{dp}_{j},14 \right),$$

Here, the maximum duration of casual partnership is 14 days whenever ${dp}_{i}$ and ${dp}_{j}$ are large.

As high *CPAR* might be due to the large number of sexual partner from group sex, so individuals with higher *CPAR* on the available list $Ca$ are more likely to engage in group sex, so in the model we set the probability whether individual $i$ can be selected for group sex in this simulation cycle as

${Pg}_{i}=1-\frac{4}{CPAR_{i}}, \mathrm{where} CPAR_{i}\geq5$.

The implementation of group sex in our model is similar with the modelling study [13] as follows. The group size has the following generalized pareto distribution

$$f\left( x \right)=\frac{1}{\sigma}\exp\left( -\frac{x-\theta}{\sigma} \right)$$

for $x>\theta$, where $\sigma=1.9, \theta=3$. These parameters are set so that the average and median group size is 4.4 and 4 respectively, matching available behavioural data [15].

The group size can be generated randomly as follows

$$X=\theta-\sigma\log\left( U\sigma\right), U\sim U\left[ 0,1 \right]$$

In the group each individual has probability of 0.5 to form a one-day casual partnership with each of other individuals in the group. Note that the individuals selected in the group do not necessarily have sexual contact with all the other members of the group.

Algorithm 3 shows the process for forming casual partnerships.

**Algorithm 3. Formation of casual partnerships**

1. For each individual $i$ in$Ca$, generate a random number $X_{i}$ from Bernoulli distribution with parameter $Pc_{i}$, if $X_{i}=1$, then put individual $i$ into $Ca_{d}.$
2. For each individual $j$ in$Ca_{d}$, generate a random number $X_{j}$ from Bernoulli distribution with parameter $Pg_{j}$, if $X_{j}=1$, then put individual $j$ into $Ca_{g}.$
3. While ( $\left| Ca_{d} \right|>1$)

Generate group size $x$;

If (form group sex with probability $1/3$^*^ and $\left| Ca_{g} \right|>x$)

Select $x$ individuals randomly from list $Ca_{g};$

Form sexual partnerships among these $x$ individuals, where each pair of these individuals has probability of 0.5 to form a one-day casual partnership;

Remove these $x$ individuals from$Ca_{g}$, $Ca_{d}$, and $Ca$.

Else

Select two individuals randomly from list $Ca_{d}$ and form casual sexual partnership;

Generate the duration of casual partnership;

Remove these two individuals from $Ca_{g}$, $Ca_{d}$, and $Ca$.

End if

End while

^*^Note that the probability of forming group sex is set to $1/3$ so that the numbers of individuals forming group sex and non-group sex casual partnership are roughly similar, where the average number in each loop is $4.4\times1/3$ and $2\times2/3$, respectively.

#### 1.2 Sexual acts and condom usage

##### Anal sex

Anal sex role preference is an important component of sexual behaviour among MSM and Table A3 shows the proportion of different preference assumed in the model. Note that if two versatile individuals form a partnership, we assume that they can be receptive or insertive during anal sex. For example, if in the model, individuals A and B are sexual partners and both versatile, during anal sex, with probability 0.5 A is insertive and B is receptive, and vice versa. Two individuals with same exclusive role preference can also form sexual partnerships but are assumed not to have anal sex during their partnership.

**Table A3 Distribution of anal sex preference**

| Role preference | Proportion | Source/Assumption |
| --- | --- | --- |
| Exclusively receptive | 11% | Adapted from a USA study by Tieu et al. [16], assuming anal sex preferences are similar in Australian MSM. |
| Exclusively insertive | 19% |  |
| Versatile | 70% |  |

The condom usage in anal sex is adapted from the GCPS Sydney 2018 [11] as shown in Table A4. In the GCPS Sydney 2018, it originally reported the condom usage with regular and casual partners, respectively, and here we adapted that information to the condom use behaviour of each individual. We assume that an individual with both types of partners would have same condom use behaviour in both types of partnerships, e.g., if he always uses a condom with casual partners, he would also always use a condom with regular partners. Then we assume that the condom usage among these who only have casual partners and that among these who have both types are the same, and equal to the condom usage with casual partners in the GCPS Sydney 2018. To have the condom usage with regular partners consistent to the reported, we adjusted the condom usage among these who only have regular partners.

**Table A4 Condom usage**

| Subgroups by partnership types | Always uses condom | Sometimes does not use a condom | Source/Assumption |
| --- | --- | --- | --- |
| Who have casual partners only | 37.12% | 62.88% | Adapted from GCPS Sydney 2018 [11] and assume condom usage are the same in these two subgroups. |
| Who have both types of partners | 37.12% | 62.88% |  |
| Who have regular partners only | 8.61% | 91.39% | Adjusted the proportion based on the population distribution. |

If an individual always using a condom encounters one sometimes not using a condom, then they would consistently use condom during their partnership, while if two individuals sometimes not using a condom form a partnership, they use condom in anal sex with probability of 0.5. Note that the efficacy of condom in preventing gonorrhoea transmission is assumed to be 87.5% (80%, 95%) according to studies [7, 17].

##### Schedule Sexual Acts

Individuals can engage in several different types of sexual act within a partnership, with probabilities governing the types of sexual act adapted from Phang *et al*. 2008 [18] and Rosenberger *et al*. 2011[19], where the porportion of males engaging each sexual practise with their most recent sexual partners was reported. These sexual acts are randomly scheduled throughout the course of the partnership according to the assumed frequencies, which was assumed by combining the assumputions in other modeling studies [6, 7] and the number of sexual acts per partner [18]. The probablities and frequencies are summarized in Table A5*.*

For example, Pharynx ↔ Pharynx (Kissing) is a common sexual act and 83% of couples would kiss each other over their partnership. In the model, we draw a random number $X$ from Bernoulli distribution with parameter$P=0.83$, and if $X=1$ the couple would kiss in the partnership, otherwise not. If a partnership would engage in Pharynx ↔ Pharynx (Kissing), they averagely kiss 2.4-3.6 times per week. Note that for casual partnerships individuals would have their prefered sexual acts at least once over their partnership.

It worth noting that any change to the assumed frequency of acts within partnerships would be compensated for by an opposing change in the estimated per-act transmission probabilities. This is because the model is calibrated to fixed prevalence targets, so any increase in one aspects of transmission is balanced by decrease in others through this process. In particular, the impact of interventions is not sensitive to changes in these sexual act frquencies. Moreover, even if the relative frequencies of the different sexual acts are changed, it is also possible to maintain the fixed calibration targets by changing the per-act transmission probabilites correspondingly.

**Table A5 Probability and frequency of sexual acts**

| Sexual Act | Values | Sources |
| --- | --- | --- |
| *Probability to engage in a sexual act* | | |
| Pharynx ↔ Pharynx (Kissing) | 0.83 | Rosenberger et al 2011 [19]. |
| Pharynx ↔ Urethra (Oral sex) | 0.825 | Phang et al 2008 [18].  Acts such as anal fingering and sharing sex toys potentially involve contacts between saliva and rectum and are also included in the *Pharynx ↔ Rectum (Rimming)* pathway, so the probability of rimming is increased accordingly |
| Pharynx ↔ Rectum (Rimming) | 0.60 |  |
| Urethra ↔ Rectum (Anal sex) | 0.478 |  |
| Urethra ↔ Urethra (Docking) | 0.03 |  |
| *Frequency of a sexual act (per week)* | | |
| Pharynx ↔ Pharynx | 2.4-3.6 | Assume to be 1.5 times of oral sex Zhang et al 2017 [20] |
| Pharynx ↔ Urethra | 1.6-2.4 | Assume to be same with anal sex, same assumption used in Hui et al [6, 21] |
| Pharynx ↔ Rectum | 1.2-1.8 | Assume based on Zhang et al 2017 [20] |
| Urethra ↔ Rectum | 1.6-2.4 | Assume based on Crawford et al 2006 [22], same assumption used in Hui et al [6, 21] and Wilson et al 2009 [13] |
| Urethra ↔ Urethra | 1.2-1.8 | Assume to be less than rimming |

### 2. Simulation approach and interventions

#### 2.1 Initial invading infection

In this study, we are particularly interested in the probability of an individual outbreak being fully contained, where outbreaks start by a single importation and the background of existing gonorrhoea strains is ignored.

When simulating the invading strain, we chose initial infectives so that across the 5000 simulations, the ratio of initial infection by site was $\text{pharynx }\text{:urethra: rectum=49:17:34}$, as estimated from site-specific incidence measured in 2015-2017 in the ACCESS database [23]. However, we adjusted this ratio slightly to account for the potential for co-infection, leading to an increased urethral proportion and decreases in the pharyngeal and rectal proportions with the revised ratio being $\text{pharynx: urethra: rectum=}\text{47:23:3}$.

The importation is then seeded at that site in a randomly selected individual who is currently infected in the equilibrium. These decisions were made in order to adjust for detection biases that occur in relation to urethral infection and to account for differences between individuals in terms of likelihood of being an index-case. Note that it is possible for the *index individual* to be infected at multiple sites.

**Algorithm 4. Introduction of Invading Strain**

1. Let  $agen{ts}_{P}, agents_{U}, agents_{R}$ be the sets of individuals who are infected at pharynx (0), urethra (1) and rectum (2), respectively.

2. Randomly generate the site of infection for index patient, where

$$site=0\left( U\leq0.47 \right)+1\left( 0.47<U\leq0.70 \right)+2\left( U>0.70 \right), U\sim U[0,1]$$

3. If $site=0$

Sample an individual from $agen{ts}_{P}$

Else if $site=1$

Sample an individual from $agents_{U}$

Else $site=2$

Sample an individual from $agents_{R}$

End if
  Denote the selected individual as$index$.

4. If $index$ individual is infected at urethra,

Set the symptom status as $symp_{U}=\left( U<0.9 \right), U\sim U[0,1]$

End if

If $index$ individual is infected at rectum,

Set the symptoms as $symp_{R}=\left( U<0.12 \right), U\sim U[0,1]$

End if

If $symp_{U}$ or $symp_{R}$ is true,

Schedule the testing and treatment within 7 days.

End if

(Note that 90% of urethral infection and 12% of rectal infection are symptomatic).

5. Except the $index$ individual, turn all other infected individuals to be susceptible with the infection and continue to run the model for 5 years.

#### 2.2 Outbreak response strategies

We consider two different patterns of testing frequency as shown in Table A6: current (CT) and recommended (RT) testing. The CT refers to the observed testing frequencies in Australian MSM and is based on data from the GCPS Sydney 2018 [11] and ACCESS [24]. For individuals who are tested fewer than once per annum, we assume their test intervals are between 360 and 720 days. The RT is based on the Australian STI Management Guidelines[25] that suggest that all MSM should test at least once a year and those at high-risk (defined as those reporting 20+ partners per year) should test every three months.

Under either scenario, for the best case, testing is assumed to occur at all anatomical sites simultaneously with 100% sensitivity, 100% treatment efficacy in individuals who test positive, and clearance of infection from all anatomical sites within 1 day. While for the worst case we assume 95% test sensitivity, 95% treatment efficacy, and 7 days clearance delay for asymptomatic infection and 1 day for symptomatic infection. Note that treatment failure only happens at the asymptomatic sites.

In Algorithm 1, in the initialization step, each individual would be given an average testing interval, and then current test dates are generated randomly. For these who have <1 test or 3+ tests, their average testing intervals are randomly sampled from $[360,720]$ or $[90,120]$, and for these who has 1, 2 or 4 tests, their average test intervals are fixed to 360, 180 or 90 days. If an individual takes a test on day $t$, his next test would be schedule at day $t+ta\times\left( 1+\frac{S}{10} \right),$ where $S\sim N(0,1)$ and $ta$ is the average test interval.

**Table A6 Annual testing frequency for current and recommended testing levels by sexual activity category**

| Testing Strategy | Testing Level by Sexual Activity Group | |
| --- | --- | --- |
|  | Low Risk^1^ | High Risk^2^ |
| Current Testing (CT) | 37.5% tested less than once  62.5% tested once | 5% tested less than once  44% tested once  22% tested twice  28% tested 3-4 times |
| Recommended Testing (RT) | 100% tested once | 100% tested 4 times |

^1^Low risk denotes individuals acquiring <20 casual partners annually and represents 73% of the modelled population.

^2^High risk denotes individuals acquiring ≥20 casual partners annually and represents 27% of the modelled population.

Contact tracing (Partner Tracing and Treating, PTT) is an important element of the clinical management of gonorrhoea cases, with benefits for both the index patients and their partners in terms of preventing re-infection and onward transmission. If contact tracing is conducted, a proportion of the index partners of patients are assumed to be tested and treated within two weeks. According to the Australia Contact Tracing Guidelines [26], gonorrhoea patients are advised to contact all sexual partners in the last two months. We consider different levels of contact tracing as follows.

a. No contact tracing.
b. Trace and treat current regular partners with different levels 80% and 100% (PTT_R80_ and PTT_R_)
c. Trace and treat current regular and a proportion of casual partners in last two months (PTT_RC_), where we considered different levels of 20%, 30%, 40% and 50%.

We assume that all current regular partners of index patients can be traced and treated, while only a proportion of casual partners can be traced and treated. For example, tracing and treating 20% casual partners (PTT_RC20_) means that a randomly selected 20% of all casual partners in the last 2 months can be traced and treated. We assume proportions of 20%, 30%, 40% or 50% for different scenarios.

We summarise the strategies combining different levels of testing and contact tracing in Table A7.

**Table A7 Outbreak response strategies**

|  | Current testing (CT) | Recommended testing (RT) |
| --- | --- | --- |
| No contact tracing | CT | RT |
| Tracing and treating 80% of regular partners (PTT_R80_) | CT+ PTT_R80_ | RT+ PTT_R80_ |
| Tracing and treating regular partners (PTT_R_) | CT+ PTT_R_ | RT+ PTT_R_ |
| Tracing and treating regular + 20% casual partners (PTT_RC20_) | CT+ PTT_RC20_ | RT+ PTT_RC20_ |
| Tracing and treating regular + 30% casual partners (PTT_RC30_) | CT+ PTT_RC30_ | RT+ PTT_RC30_ |
| Tracing and treating regular + 40% casual partners (PTT_RC40_) | CT+ PTT_RC40_ | RT+ PTT_RC40_ |
| Tracing and treating regular + 50% casual partners (PTT_RC50_) | CT+ PTT_RC50_ | RT+ PTT_RC50_ |
| Response with 6-month delay post the first detection | CT+PTT_R (6-months)_ | RT _(6-months)_ |
| Recommended testing frequency by STIGMA guideline | None | RT_STIGMA_ |
| 100% condom use for anal sex | CT+C_A100_ | None |
| 100% condom use for both oral and anal sex | CT+C_AO100_ | None |

#### 2.3 Model Calibration

##### Calibration targets

The model was calibrated to estimated site-specific prevalence among overall Australian MSM [20] as shown in the Table A8. We note that these values reflect the expected prevalence observed in a community prevalence survey with urethral prevalence much lower than as measured in clinic populations, where attendance for treatment of urethral infection is a significant source of bias. Note that we also considered calibration to site-specific incidence as measured in ACCESS [ref] but found this led to lower prevalence values than shown in Table A8, and more optimistic results for the effect of interventions. Therefore, we opted to calibrate to the prevalence, where our evaluation of impact was more conservative.

We use $(prev_{PH},prev_{UR}, prev_{RE})$ to represent the target values of site-specific prevalence.

**Table A8 Calibration targets of site-specific prevalence**

| Sites | Values | Data sources |
| --- | --- | --- |
| Pharynx $prev_{PH}$ | 8.6% (7.7-9.5%) | These values were estimated by Zhang *et al.* [20], and the prevalence levels were comparable with studies [27, 28], where high prevalence of pharyngeal and rectal infection and low prevalence of urethral infection were reported. |
| Urethra $prev_{UR}$ | 0.2% (0.04-3.5%) |  |
| Rectum $prev_{RE}$ | 8.3% (7.4-9.1%) |  |

The objective function in this case is highly stochastic, multimodal and computationally expensive to evaluate and therefore the optimal solution is difficult to find. The Nelder-Mead method is not suitable for this problem due to a large number of local minima, while the Cross-entropy method [29] was also tested but failed to shrink ranges parameter space effectively. Therefore, we adopted a relatively simple approach in which we first defined feasible parameter ranges for the transmission probabilities based on other modelling studies and prior knowledge [6, 7] as shown in Table A9 below.

**Table A9 Calibrated per-act transmission probabilities**

| **Parameter** | **Transmission direction** | **Parameter boundaries** | **Calibrated** **solution (best)** | **Calibrated** **solution (worst)** |
| --- | --- | --- | --- | --- |
| $p_{1}$ | Pharynx → Pharynx | (0,0.2) | 0.0978 | 0.0847 |
| $p_{2}$ | Pharynx →urethra | (0,0.04) | 0.0098 | 0.0094 |
| $p_{3}$ | Urethra →pharynx | (0.2,0.8) | 0.4379 | 0.4690 |
| $p_{4}$ | Pharynx →rectum | (0,0.3) | 0.0763 | 0.0589 |
| $p_{5}$ | Rectum →pharynx | (0,0.5) | 0.0031 | 0.0049 |
| $p_{6}$ | Urethra →rectum | (0.2,0.9) | 0.6339 | 0.6940 |
| $p_{7}$ | Rectum →urethra | (0,0.1) | 0.0188 | 0.0125 |
| $p_{8}$ | Urethra →urethra | (0,0.01) | 0.0048 | 0.0042 |

Treating these ranges as uniform distributions for the transmission probabilities, we then generated 10,000 sampled parameter sets, which were compared against the prevalence values using the objective function described below. The average objective function over 5 runs for each parameter set was then calculated and the simulation with the smallest average was then used for the baseline results and analysis of interventions.

##### Objective functions

We considered 5 types of sexual practices in the model, resulting in 8 transmission routes of infection: pharynx to pharynx, pharynx to urethra, urethra to pharynx, pharynx to rectum, rectum to pharynx, urethra to rectum, rectum to urethra, and urethra to urethra. We denote the per-act transmission probabilities of each route as${(p}_{1},...,p_{8})$, which are the parameters to be estimated. Note that the only parameters to be obtained from calibration process are ${(p}_{1},...,p_{8})$, and all other parameters are randomly sampled from their fixed range during model running and not to be calibrated.

The transmission probabilities are varied so that the model-generated site-specific prevalence at equilibrium matches the calibration targets. In detail, if given a set of transmission probabilities, we initialize sexual partnership network of the model and run the model for 5 years before introducing the background strains. After another 5 years of model running when the prevalence gets to stable level, we generate and record the quarterly average site-specific prevalence for the next 30 years, with which we can compute the squared error between simulated site-specific prevalence and targeted prevalence. Then we have the squared error function as follows that depends on the per-act transmission probabilities.

$$f\left( p_{1},\ldots,p_{8} \right)=\frac{1}{T_{prev}}\sum_{t=1}^{T_{prev}} \left( prev_{PH,t}-prev_{PH} \right)^{2}+\left( prev_{UR,t}-prev_{UR} \right)^{2}+\left( prev_{RE,t}-prev_{RE} \right)^{2},$$

$s.t. 0\leq p_{i}\leq1, i=1,\ldots, 8$.

where $(prev_{PH,t},prev_{UR,t}, prev_{RE,t})$ represent the average daily prevalence per quarter for pharyngeal, urethral and rectal infection respectively, and $T_{prev}$ is the total number of quarterly periods (i.e $T_{prev}=120$). For example, $(prev_{PH,2}, prev_{UR,2}, prev_{RE,2})$ is the average daily prevalence from 4^th^ to 6^th^ month, which is the second calculation period.

The per-act transmission probabilities can be obtained by minimizing the squared error functions. As the objective functions $f\left( p_{1},...,p_{8} \right)$ are highly stochastic, for each realization of parameters $\left( p_{1},...,p_{8} \right),$ we run the model for 5 times and use the average as the objective value. The process is summarized in Algorithm 4.

**Algorithm 4.** Calibration with random sampling

1 Initialize boundaries of parameter space

Set boundaries of parameter space based on the results in [6, 7].

2 Sample and calculate the objective function.

Randomly and uniformly draw 10,000 samples from the parameter space, denoted as$P_{1}, P_{2},\ldots,P_{10000}$.

For each sample $P_{i}$, we calculate objective functions and have $\left( P_{i},f_{i} \right),$

$for i=1,\ldots, 10,000$.

3 Summarize results.

Rank the samples based on the objective value and select the best solution as the estimation of parameters.

### 3. Results

In this section, we will present some model verification about sexual partnerships and infection transmission, and additional results about responses strategies.

#### 3.1 Additional calibration and validation results

##### Output of Sexual partnership

We run the sexual partnership network for 10 years and repeat 20 times. Figure A2 shows the proportion of the population in each of partner preference categories by year. Note that the proportions denoted ‘Single’ are those individuals who have neither casual nor regular partners during a given year but individuals represented in this group may have partners in other years. With equilibrium reached at around year 3, this shows that the 5 years burn-in period for the sexual partnership network as described in Algorithm 1 is sufficient.

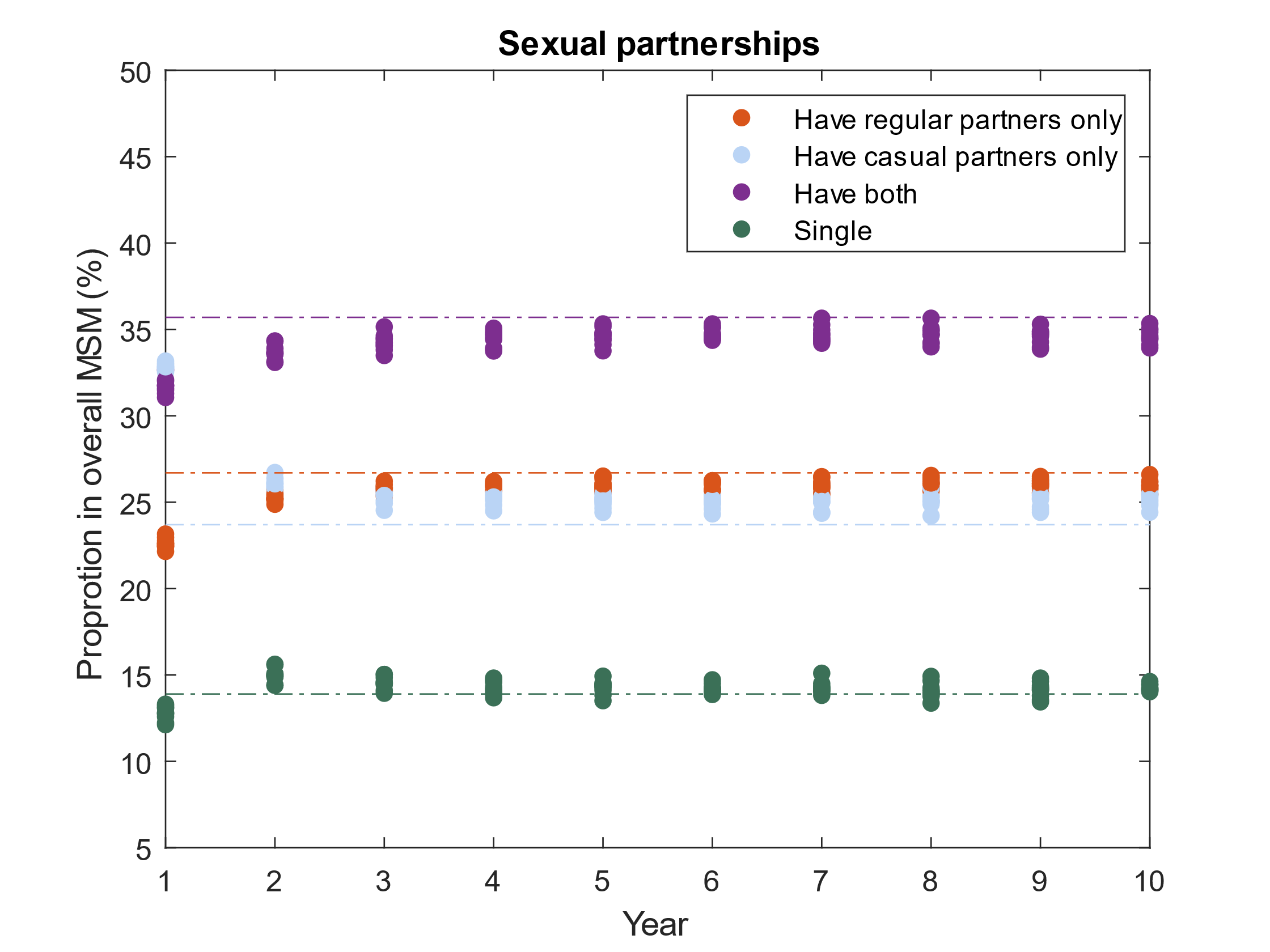

Figure A2. Distribution of population by different sexual partner preference in each year. The flat dashed lines are the data from GCPS Sydney 2018

Figure A3 presents compares the simulated distribution of casual partners acquisition rates with those reported in the HIM study [13, 22]. The simulated rates were calculated by running the model for 10 years and summarising casual partner numbers per individual for every 6 month period. These frequencies were averaged over each of the 20 6-month periods and then this process was then repeated 20 times to account for stochastic variation with the final distribution representing an average over these 20 simulations. The model output is generally close to the HIM study data, with a slight excess of individuals with 2-5 partnerships, while those with 50+ partnerships are underestimated.

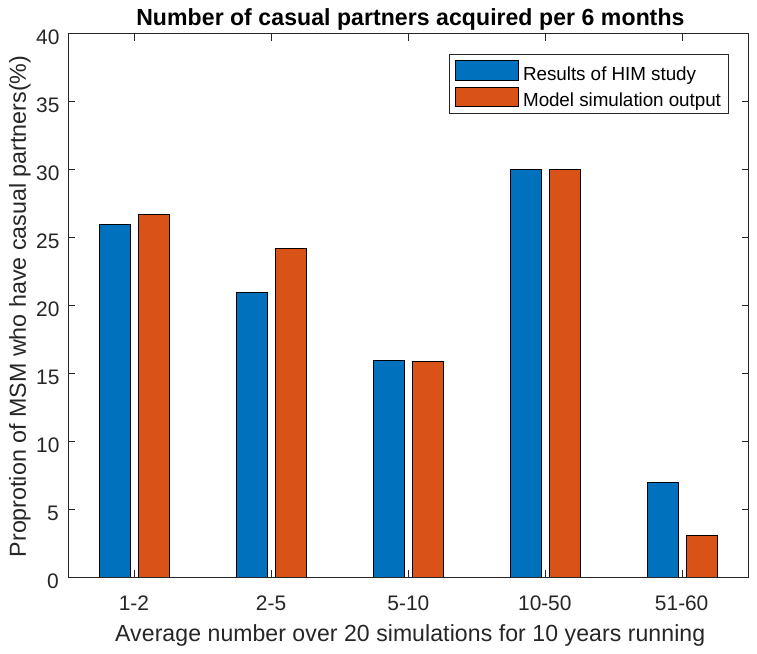

Figure A3 Comparison of number of casual partners acquired per 6 months: model output .vs HIM study

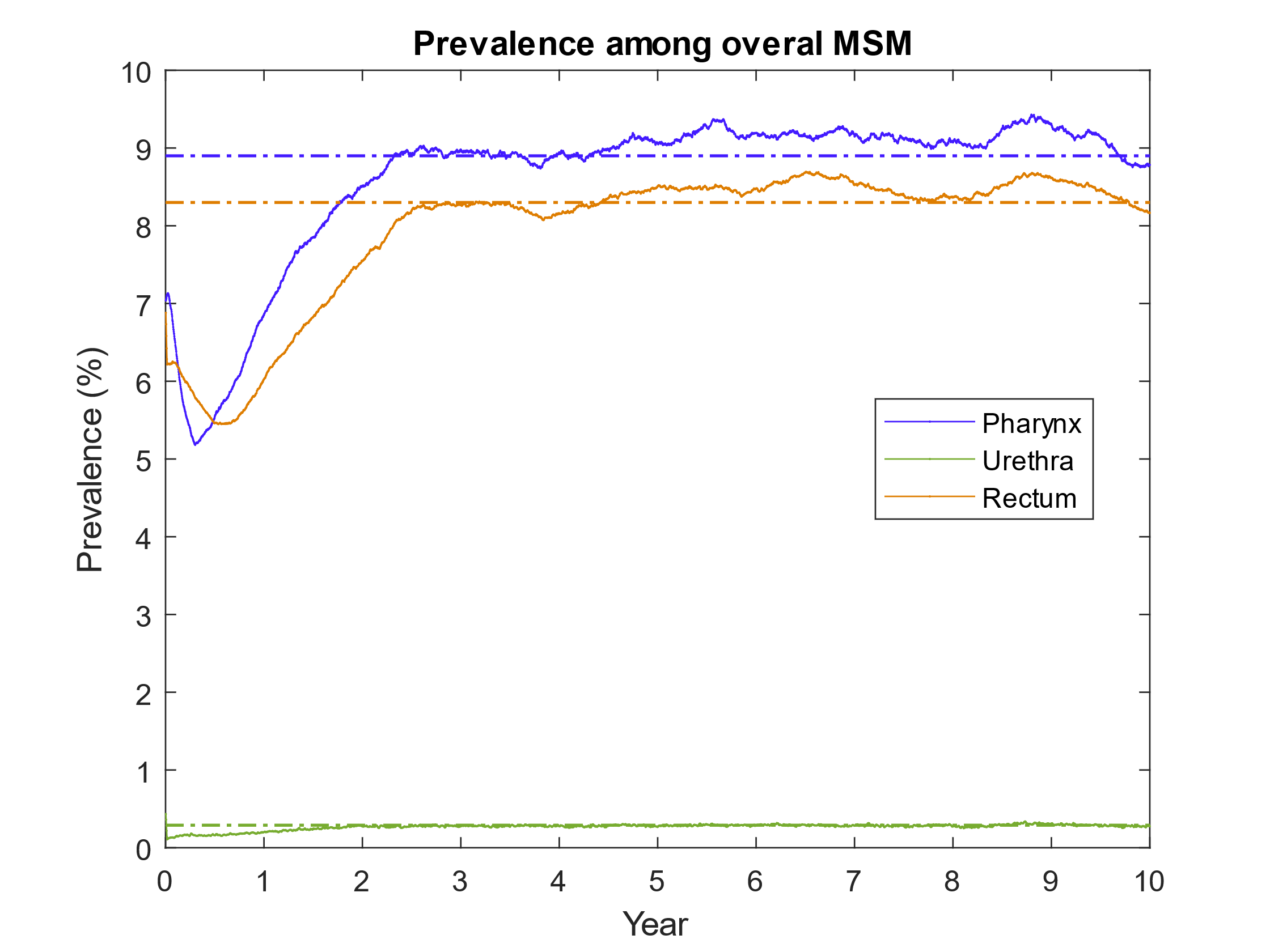

Figure A4 Site-specific prevalence among overall MSM

##### Output of site-specific prevalence

Figure A4 shows the evolution of site-specific prevalence in the first 10 years after introduction of the background infection, which is introduced after first running the sexual partnership network simulation for 5 years without infection. As we can see, the simulated prevalence reaches the equilibrium states at 3 years post introduction, which indicates that 5 years used in the Algorithm 1 are enough for infection transmission reaching the equilibrium.

#### 3.2 Scenario and sensitivity analysis

##### Additional results under base case

In Table A10, the proportions of the 5000 simulations for each scenario are broken down by persistence and detection status at 6 months and 2 and 5 years post importation under base case. This table also includes additional scenarios, with CT+C_A100_ referring to 100% condom use for anal sex, CT+C_AO100_ for increasing to 100% condom use for both anal and oral sex and CT+ PTT_R (6-months)_ and RT _(6-months)_ incorporating a 6 month delay in applying PTT_R_ and RT, while RT_STIGMA_ refers to increasing recommended testing to conform with 2019 STIGMA guidance.

**Table A10 Persistence probabilities at the 6 months and 2 and 5 years post importation (Base)**

| Response strategies | Detected and persisting simulations (%) | | | Extinct simulations (%) | | |
| --- | --- | --- | --- | --- | --- | --- |
|  | 6 months | 2 years | 5 years | 6 months | 2 year | 5 year |
| CT | 25.8 | 15.8 | 14.0 | 66.3 | 84.2 | 86.0 |
| CT+C_A100_ | 25.8 | 15.8 | 13.8 | 66.3 | 84.2 | 86.2 |
| CT+C_AO100_ | 25.4 | 14.8 | 12.7 | 66.7 | 85.2 | 87.3 |
| CT+PTT_R80_ | 18.1 | 7.10 | 4.46 | 74.1 | 92.9 | 95.5 |
| CT+ PTT_R_ | 15.5 | 4.3 | 1.44 | 76.7 | 95.7 | 98.6 |
| CT+ PTT_R (6-months)_ | 25.8 | 12.4 | 6.34 | 66.3 | 87.6 | 93.6 |
| RT | 25.5 | 13.9 | 9.8 | 66.6 | 86.1 | 90.2 |
| RT _(6-months)_ | 25.8 | 15.5 | 12.0 | 66.3 | 84.5 | 87.9 |
| RT_STIGMA_ | 25.4 | 13.1 | 8.4 | 66.7 | 86.9 | 91.6 |
| RT+PTT_R80_ | 17.8 | 3.98 | 0.02 | 74.3 | 96.0 | 100 |
| RT +PTT_R_ | 15.5 | 1.34 | 0 | 76.6 | 98.7 | 100 |
| CT+ PTT_RC20_ | 14.4 | 2.94 | 0.24 | 77.7 | 97.1 | 99.8 |
| CT+ PTT_RC30_ | 13.7 | 2.28 | 0.1 | 78.4 | 97.7 | 99.9 |
| CT+ PTT_RC40_ | 12.7 | 1.9 | 0.12 | 79.4 | 98.1 | 99.9 |
| CT+ PTT_RC50_ | 12.3 | 1.36 | 0 | 79.8 | 98.6 | 100 |
| RT+ PTT_RC20_ | 14.3 | 0.78 | 0 | 77.8 | 99.2 | 100 |
| RT+ PTT_RC30_ | 13.4 | 0.44 | 0 | 78.7 | 99.6 | 100 |
| RT+ PTT_RC40_ | 12.7 | 0.3 | 0 | 79.4 | 99.7 | 100 |
| RT+ PTT_RC50_ | 12.2 | 0.18 | 0 | 79.9 | 99.8 | 100 |
| **Note that the proportion of undetected and persisting simulations (%) was 7.88% at the 6 months post importation under all scenarios and 0% at other time points.** | | | | | | |

Under base-case assumptions, universal condom-use for anal sex is of limited impact, while adding universal condom-use for oral sex has only a similar impact to moving to recommended treatment. Figure A5 shows the persistence probability over time post importation for the 8 additional response strategies introduced in Table A10.

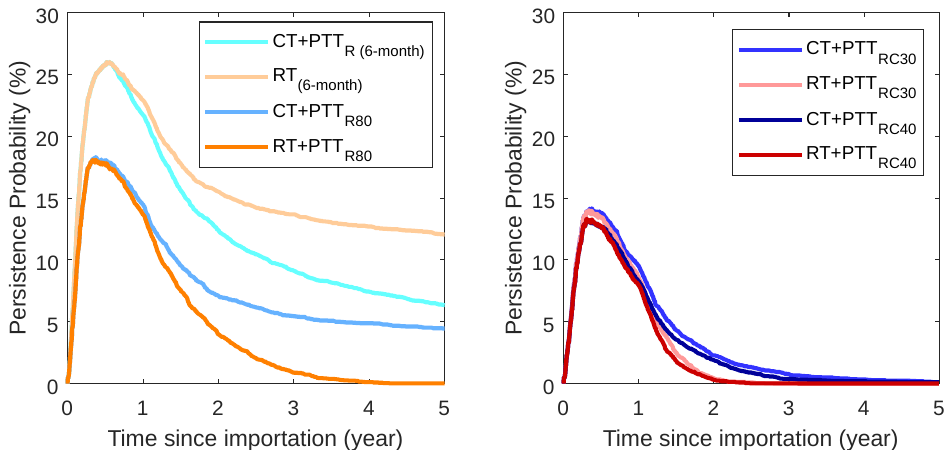

*Figure A5. The persistence probability as a function of time from importation of NG strain and response strategies.*

Figure A6 shows the effects of these additional strategies on outbreak size and the probability of eliminating the invading strain over the first two years after detection.

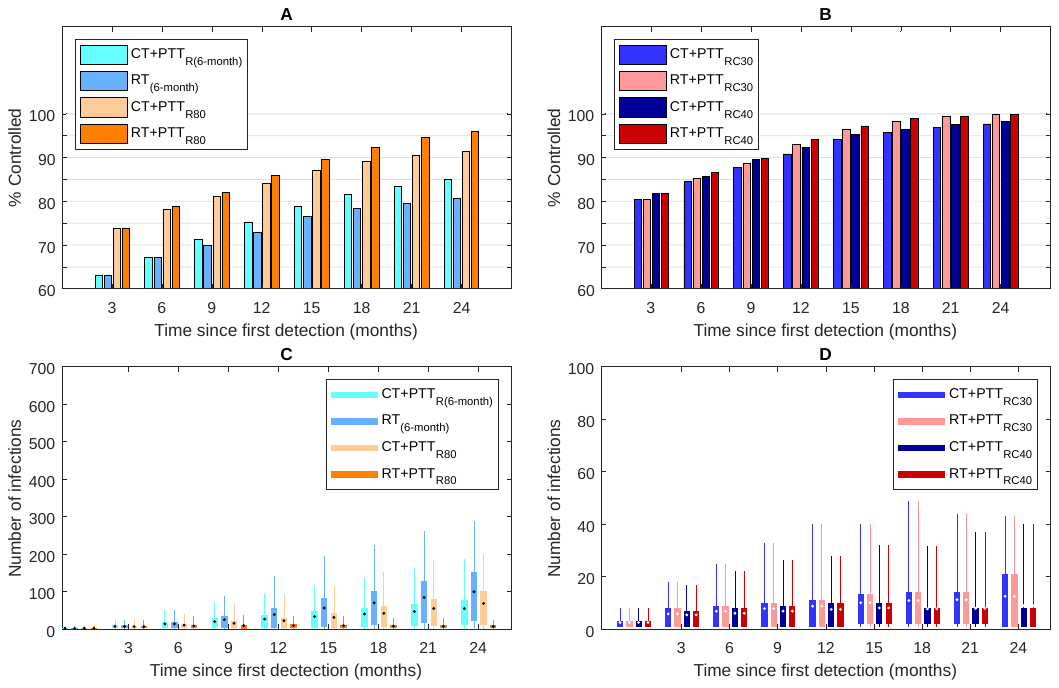

Figure A6. Panel A and B show the proportion of those simulations in which the invading strain becomes extinct, as a function of the time from the first detection and outbreak response strategy. Panel C and D shows the outbreak size of invading NG strain among non-extinct simulations as a function of time from the first detection and outbreak response strategy. The box denotes the interquartile range (25% to 75%), the whiskers the quantile (5% to 95%), the horizontal line in each box the median, and the dot in each box the mean. Outbreak size is the number or infected individuals at each time point for simulations in which the invading strain persists.

##### Results under more pessimistic treatment assumptions

We also consider a more pessimistic set of assumptions for treatment where we assume 95% test sensitivity, 95% treatment efficacy, and a 7-day clearance delay for asymptomatic infection. In Table A11 and Figure A7, we present the results of simulations for the model after recalibration using the above assumptions.

**Table A11 Persistence probabilities at the 6 months and 2 and 5 years post importation (Worst)**

| Response strategies | Detected and persisting simulations (%) | | | Extinct simulations (%) | | |
| --- | --- | --- | --- | --- | --- | --- |
|  | 6 months | 2 years | 5 years | 6 months | 2 year | 5 year |
| CT | 30.7 | 18.3 | 15.4 | 60.7 | 81.7 | 84.6 |
| CT+PTT_R80_ | 22.3 | 8.5 | 4.9 | 69.2 | 91.5 | 95.1 |
| CT+ PTT_R_ | 20.2 | 6.9 | 2.9 | 71.3 | 93.1 | 97.1 |
| RT | 30.1 | 15.5 | 8.0 | 61.3 | 84.5 | 91.9 |
| RT_STIGMA_ | 30.0 | 13.6 | 5.6 | 61.5 | 86.4 | 94.4 |
| RT+PTT_R80_ | 22.1 | 4.9 | 0.1 | 69.3 | 95.1 | 99.9 |
| RT +PTT_R_ | 20 | 3.6 | 0.04 | 71.4 | 96.4 | 100 |
| CT+ PTT_RC20_ | 20.2 | 5.9 | 1.8 | 71.3 | 94.1 | 98.2 |
| CT+ PTT_RC30_ | 19.6 | 5.8 | 1.4 | 71.8 | 94.2 | 98.6 |
| CT+ PTT_RC40_ | 18.5 | 4.6 | 0.9 | 72.9 | 95.4 | 99.1 |
| CT+ PTT_RC50_ | 18.1 | 4.2 | 0.4 | 73.4 | 95.8 | 99.6 |
| RT+ PTT_RC20_ | 19.5 | 2.5 | 0 | 71.9 | 97.5 | 100 |
| RT+ PTT_RC30_ | 18.9 | 1.8 | 0 | 72.5 | 98.2 | 100 |
| RT+ PTT_RC40_ | 18.0 | 1.8 | 0 | 73.4 | 98.2 | 100 |
| RT+ PTT_RC50_ | 17.6 | 1.7 | 0 | 73.8 | 98.3 | 100 |
| **Note that the proportion of undetected and persisting simulations (%) was 8.6% at the 6 months post importation under all scenarios and 0% at other time points.** | | | | | | |

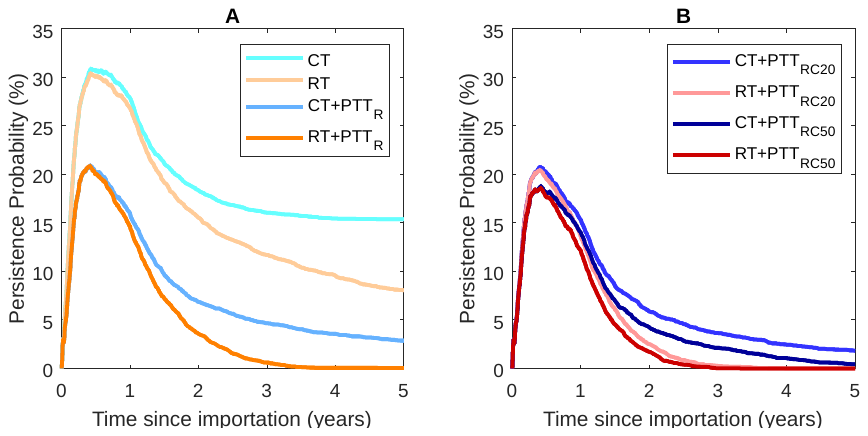

*Figure A7. The persistence probability as a function of time from importation of NG strain and response strategies.*

Outbreak sizes for these more pessimistic treatment assumptions are captured in Figure A8. Due to the recalibration leading to somewhat lower per-act transmission probabilities, these more pessimistic assumptions do not necessarily lead to larger outbreak sizes as shown in Panels C and D.

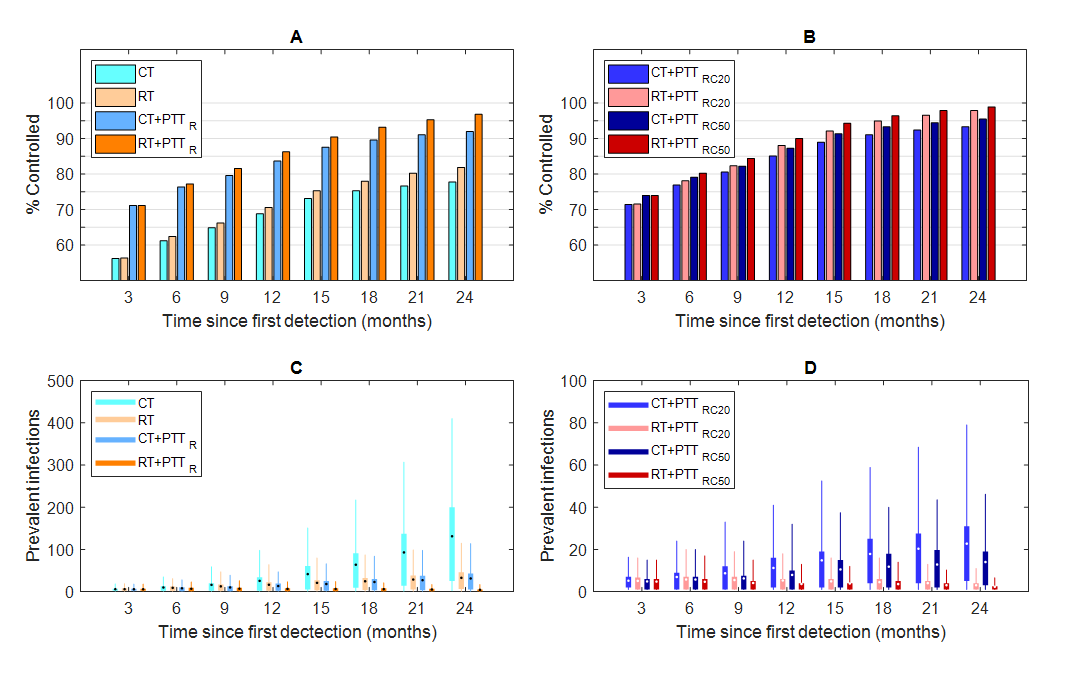

*Figure A8. Panel A and B show the proportion of those simulations in which the invading strain becomes extinct, as a function of the time from the first detection and outbreak response strategy. Panel C and D shows the outbreak size of invading NG strain among non-extinct simulations as a function of time from the first detection and outbreak response strategy. The box denotes the interquartile range (25% to 75%), the whiskers the quantile (5% to 95%), the horizontal line in each box the median, and the dot in each box the mean. Outbreak size is the number or infected individuals at each time point for simulations in which the invading strain persists.*

Table A12 shows this effect of recalibration on outbreak sizes in more detail over the 5 year simulation period, with only current and recommended test levels leading to higher prevalence under the base-case assumptions than with more pessimistic treatment assumptions (highlighted rows).

**Table A12 Comparison of prevalence % at the 2 and 5 years post importation (Best and Worst)**

|  | The base-case | | The worst case | |
| --- | --- | --- | --- | --- |
|  | 2 years | 5 years | 2 years | 5 years |
| CT | 2.34(0.01,11.21) | 14.77(0.12,19.25) | 0.99(0.01,7.71) | 9.21 (0.01,15.21) |
| CT+ PTT_R_ | 0.25(0.01,1.27) | 0.46(1,2.23) | 0.28(0.01,1.87) | 0.66(0.01,2.30) |
| CT+ PTT_RC20_ | 0.15(0.01,1.01) | 0.27(0.01,1.13) | 0.20(0.01,1.39) | 0.35(0.01,1.80) |
| CT+ PTT_RC50_ | 0.07(0.01,0.45) | 0.00(0,0) | 0.13(0.01,1.01) | 0.13(0.01,0.35) |
| RT | 0.58(0.01,3.63) | 1.90(0.01,6.19) | 0.29(0.01,2.43) | 0.60(0.01,2.55) |
| RT+ PTT_R_ | 0.04(0.01,0.17) | 0.00(0,0) | 0.05(0.01,0.24) | 0.01(0.01, 0.01) |
| RT+ PTT_RC20_ | 0.03 (0.01,0.12) | 0.00(0,0) | 0.04(0.01,0.18) | 0.00(0,0) |
| RT+ PTT_RC50_ | 0.01(0.01, 0.03) | 0.00(0,0) | 0.03(0.01,0.16) | 0.00(0,0) |
| CT+ PTT_R (6-months)_ | 0.48(0.01,3.32) | 1.05(0.01,4.96) |  |  |
| RT _(6-months)_ | 0.86(0.01,5.42) | 2.25 (0.01,6.32) |  |  |

##### Effect of a higher proportion of asymptomatic urethral infections

Table A13 shows the results under the assumption that 40% of urethral infection are asymptomatic. Under this scenario, recalibration again leads to lower per-act transmission probabilities and stronger effects of control strategies. In particular, strategies involved increased condom-use are much more effective with this level of asymptomatic urethral infection.

**Table A13 Persistence probabilities at the 6 months and 2 and 5 years post importation (U)**

| Response strategies | Detected and persisting simulations (%) | | | Extinct simulations (%) | | |
| --- | --- | --- | --- | --- | --- | --- |
|  | 6 months | 2 years | 5 years | 6 months | 2 year | 5 year |
| CT | 22.9 | 11.0 | 8.7 | 68.8 | 89.0 | 91.3 |
| CT+C_A100_ | 22.7 | 9.1 | 5.7 | 69.0 | 90.9 | 94.3 |
| CT+C_AO100_ | 21.9 | 4.9 | 0.2 | 69.8 | 95.1 | 99.8 |
| CT+ PTT_R_ | 13.1 | 3.2 | 1.2 | 78.6 | 96.8 | 98.8 |
| RT | 22.2 | 8.3 | 2.1 | 69.5 | 91.7 | 97.9 |
| RT +PTT_R_ | 12.6 | 1.3 | 0 | 79.1 | 98.7 | 100 |

**Note that the proportion of undetected and persisting simulations (%) was 8.3% at the 6 months post importation under all scenarios and 0% at other time points.**

References

1. Rebe K, Lewis D, Myer L, et al. A Cross Sectional Analysis of Gonococcal and Chlamydial Infections among Men-Who-Have-Sex-with-Men in Cape Town, South Africa. PLOS ONE **2015**; 10:e0138315.

2. Ryder N, Lockart IG, Bourne C. Is screening asymptomatic men who have sex with men for urethral gonorrhoea worthwhile? Sexual Health **2010**; 7:90-1.

3. Dudareva-Vizule S, Haar K, Sailer A, et al. Prevalence of pharyngeal and rectal Chlamydia trachomatis and Neisseria gonorrhoeae infections among men who have sex with men in Germany. Sex Transm Infect **2013**:sextrans-2012-050929.

4. Chow EP, Camilleri S, Ward C, et al. Duration of gonorrhoea and chlamydia infection at the pharynx and rectum among men who have sex with men: a systematic review. Sex Health **2016**; 13:199-204.

5. Fairley CK, Chen MY, Bradshaw CS, Tabrizi SN. Is it time to move to nucleic acid amplification tests screening for pharyngeal and rectal gonorrhoea in men who have sex with men to improve gonorrhoea control? Sex Health **2011**; 8:9-11.

6. Hui B, Fairley C, Chen M, et al. Oral and anal sex are key to sustaining gonorrhoea at endemic levels in MSM populations: a mathematical model. Sex Transm Infect **2015**; 91:365-9.

7. Zhang L, Regan DG, Chow EP, et al. Neisseria gonorrhoeae transmission among men who have sex with men: an anatomical site-specific mathematical model evaluating the potential preventive impact of mouthwash. Sex Transm Dis **2017**; 44:586-92.

8. Fairley CK, Chow EPF, Hocking JS. Early presentation of symptomatic individuals is critical in controlling sexually transmissible infections. Sexual Health **2015**; 12:181-2.

9. Harrison WO, Hooper RR, Wiesner PJ, et al. A trial of minocycline given after exposure to prevent gonorrhea. N Engl J Med **1979**; 300:1074-8.

10. Chow EP, Read TR, Wigan R, et al. Ongoing decline in genital warts among young heterosexuals 7 years after the Australian human papillomavirus (HPV) vaccination programme. Sex Transm Infect **2014**:sextrans-2014-051813.

11. Broady T, Mao, L., Lee, E., Bavinton, B., Keen, P., Bambridge, C., Mackie, B., Duck, T., Cooper, C., Prestage, G., & Holt, M. . Gay Community Periodic Survey: Sydney 2018. Centre for Social Research in Health, UNSW Sydney, **2018**.

12. Prestage GP, J. H, J. B, al. e. TOMS-Three or More Study. Sydney: National Centre in HIV Epidemiology and Clinical Research. University of New South Wales **2008**.

13. Wilson D, Prestage G, Donovan B, Gray R, Hoare A, McCann P. Phase A of the National Gay Men’s Syphilis Action Plan: modelling evidence and research on acceptability of interventions for controlling syphilis in Australia. Sydney: National Centre in HIV Epidemiology and Clinical Research **2009**.

14. Fogarty A, Mao L, Zablotska I, et al. The Health in Men and Positive Health cohorts A comparison of trends in the health and sexual behaviour of HIV-negative and HIV-positive gay men, 2002-2005. **2006**.

15. Prestage GP, Hudson J, Down I, et al. Gay men who engage in group sex are at increased risk of HIV infection and onward transmission. AIDS and Behavior **2009**; 13:724.

16. Tieu HV, Li X, Donnell D, et al. Anal sex role segregation and versatility among men who have sex with men: EXPLORE Study. J Acquir Immune Defic Syndr **2013**; 64:121-5.

17. Warner L, Newman DR, Austin HD, et al. Condom Effectiveness for Reducing Transmission of Gonorrhea and Chlamydia: The Importance of Assessing Partner Infection Status. Am J Epidemiol **2004**; 159:242-51.

18. Phang CW, Hocking J, Fairley CK, Bradshaw C, Hayes P, Chen MY. More than just anal sex: the potential for STI transmission among men visiting sex on premises venues. Sex Transm Infect **2008**.

19. Rosenberger JG, Reece M, Schick V, et al. Sexual behaviors and situational characteristics of most recent male-partnered sexual event among gay and bisexually identified men in the United States. J Sex Med **2011**; 8:3040-50.

20. Zhang L, Regan DG, Chow EP, et al. Neisseria gonorrhoeae transmission among men who have sex with men: an anatomical site-specific mathematical model evaluating the potential preventive impact of mouthwash. Sex Transm Dis **2017**; 44:586-92.

21. Hui BB, Whiley DM, Donovan B, Law MG, Regan DG. Identifying factors that lead to the persistence of imported gonorrhoeae strains: a modelling study. Sex Transm Infect **2016**:sextrans-2016-052738.

22. Crawford JM, Kippax SC, Mao L, et al. Number of risk acts by relationship status and partner serostatus: findings from the HIM cohort of homosexually active men in Sydney, Australia. AIDS Behav **2006**; 10:325-31.

23. Callander D, Guy R, Fairley CK, et al. Gonorrhoea gone wild: rising incidence of gonorrhoea and associated risk factors among gay and bisexual men attending Australian sexual health clinics. Sex Health **2019**; 16:457-63.

24. Callander D, Donovan B, Guy R. The Australian Collaboration for Coordinated Enhanced Sentinel Surveillance of Sexually Transmissible Infections and Blood Borne Viruses: NSW STI Report 2007-2014. . Sydney, NSW: UNSW Australia., **2015**.

25. Simms KT, Laprise JF, Smith MA, et al. Cost-effectiveness of the next generation nonavalent human papillomavirus vaccine in the context of primary human papillomavirus screening in Australia: a comparative modelling analysis. Lancet Public Health **2016**; 1:e66-e75.

26. Fraser C, Tomassini JE, Xi L, et al. Modeling the long-term antibody response of a human papillomavirus (HPV) virus-like particle (VLP) type 16 prophylactic vaccine. Vaccine **2007**; 25:4324-33.

27. Cornelisse VJ, Chow EP, Huffam S, et al. Increased Detection of Pharyngeal and Rectal Gonorrhea in Men Who Have Sex With Men After Transition From Culture To Nucleic Acid Amplification Testing. Sex Transm Dis **2017**; 44:114-7.

28. Fairley CK, Hocking JS, Zhang L, Chow EP. Frequent transmission of gonorrhea in men who have sex with men. Emerg Infect Dis **2017**; 23:102.

29. Benham T, Duan Q, Kroese DP, Liquet B. CEoptim: Cross-Entropy R Package for Optimization. J Stat Softw **2017**; 76:29.
